# A Phenome-wide association study of genetically determined nicotine metabolism reveals novel links with health-related outcomes

**DOI:** 10.1101/2023.12.22.23300430

**Authors:** Jadwiga Buchwald, Terho Lehtimäki, Olli Raitakari, Veikko Salomaa, Jaakko Kaprio, Matti Pirinen

## Abstract

**Background:** Faster nicotine metabolism, defined as the nicotine metabolite ratio (NMR), is known to associate with heavier smoking and challenges in smoking cessation. However, the broader health implications of genetically determined nicotine metabolism are not well characterized.

**Methods:** We performed a hypothesis-free phenome-wide association study (PheWAS) of over 21,000 outcome variables from UK Biobank (UKB) to explore how the NMR (measured as the 3-hydroxycotinine-to-cotinine ratio) associates with the phenome. As the exposure variable, we used a genetic score for faster nicotine metabolism based on 10 putative causal genetic variants, explaining 33.8 % of the variance in the NMR. We analysed ever and never smokers separately to assess whether a causal pathway through nicotine metabolism is plausible.

**Results:** A total of 57 outcome variables reached phenome-wide significance at a false discovery rate of 5 %. We observed expected associations with several phenotypes related to smoking and nicotine, but could not replicate prior findings on cessation. Importantly, we found novel associations between genetically determined faster nicotine metabolism and adverse health outcomes, including unfavourable liver enzyme and lipid values, as well as increased caffeine consumption. These associations did not appear to differ between ever and never smokers, suggesting the corresponding pathways may not involve nicotine metabolism. No favourable health outcomes were linked to genetically determined faster nicotine metabolism.

**Conclusions:** Our findings support a possibility that a future smoking cessation therapy converting fast metabolizers of nicotine to slower ones could work without adverse side effects and potentially even provide other health-related benefits.

## Introduction

Smoking remains a leading cause of global morbidity and mortality [1]. While global smoking prevalence has declined due to effective tobacco control policies, the prevalence has risen or remained stagnant in many countries [2]. Furthermore, the emergence of new nicotine products, such as e-cigarettes, which have aggressively been targeted at the youth, has created a whole new generation of addicted individuals in some countries [2, 3]. Nicotine, a toxic substance in itself [4, 5], is as addictive as cocaine and heroine [6].

The Nicotine Metabolite Ratio, NMR, measured as the 3-hydoxycotinine-to-cotinine ratio (3HC/Cot), is an established biomarker for the rate of nicotine metabolism [7]. Individuals with higher NMR values, reflecting faster nicotine metabolism, typically smoke more [8, 9] and find quitting more challenging [10–12]. Personalizing cessation treatments based on an individual’s NMR could improve cessation rates [13]. Another approach would be to develop smoking cessation drugs tailored to modulate metabolization rates.

The NMR is highly heritable [14], and understanding which genetic loci it associates with, can illuminate potential drug targets. The largest genome-wide association study (GWAS) on the NMR to date identified two independent association loci on chromosomes 4 and 19, explaining 1.8 % and 36.4 % of the variation in the NMR, respectively [15]. Notably, the chromosome 19 locus contains *CYP2A6*, which codes for CYP2A6, the main metabolic enzyme for nicotine [16]. Nicotine is metabolized primarily to cotinine (Cot) (up to 75 %), mainly by CYP2A6, and Cot is metabolized primarily to 3-hydroxycotinine (3HC) (up to 40 %), exclusively by CYP2A6 [16]. Thus, their ratio, 3HC/Cot, i.e. the NMR, also reflects CYP2A6 activity [7]. A smaller fraction of nicotine and cotinine is metabolized through other enzymatic pathways [16].

It is also essential to understand what traits or diseases the NMR is associated with. This knowledge is critical for assessing possible side-effects and other opportunities for drug development. A previous study by Chenowth et al. showed the NMR to be associated with ethnicity, gender, hormonal replacement therapy, BMI, cigarettes smoked per day (CPD), and number of alcohol drinks/week [17]. In that hypothesis-driven study, the variables investigated had been carefully chosen based on previous literature. Therefore, unknown and potentially important associations may have been missed.

Phenome-wide association studies (PheWASs) present a hypothesis-free approach to discover novel associations [18]. PheWASs aim to identify associations with a genetic instrument (e.g. a single nucleotide polymorphism (SNP) or genetic risk score), proxying a given variable of interest, across an array of phenotypes (the phenome). Two PheWASs of CYP2A6 activity have been published to date. The first, assessed 358 traits for nine *CYP2A6* SNPs, and identified an association between one of the SNPs and hearing loss among the nicotine-exposed subgroup but not among the nicotine-unexposed subgroup [19]. The second, was a PheWAS of a genetic score for CYP2A6 activity in UKB [20]. However, they limited their analyses to a total of 1,029 disease endpoints, based on ICD-9 and ICD-10 diagnostic rubrics. They found associations with lung cancer and other known smoking related diseases; no associations were seen among their subsets of former or never smokers.

Our aim was to use the UKB data (N = 343,662), without limiting ourselves to any specific category of phenotypes, to assess how the NMR is associated with the phenome. This is the largest PheWAS on the NMR to date, encompassing over 21,000 outcome variables. Our study is also the first to explicitly focus on the NMR, rather than solely on CYP2A6 activity.

We created a genetic score for the NMR from ten putative causal SNPs, explaining 33.8 % of the variance in the NMR. We used the sofware package PHESANT [21] which enabled us to incorporate all variable types (continuous, binary, categorical, ordinal) in the PheWAS, and thus uncover novel associations. Importantly, we used the GxE MR-pheWAS approach [22], meaning that we ran the PheWAS also separately for ever and never smokers. The approach permitted us to distinguish whether the associations reflected a causal pathway through a) the NMR, either directly or through other traits such as amount smoked (effect only seen in ever smokers), b) some other pathway not including the NMR (same effect seen also in never smokers), or c) both (effect only seen in never smokers, or there are quantitative or directional differences in the effect sizes between ever and never smokers). Our findings contribute valuable information for drug development and personalized interventions for treating nicotine addiction, as well as, for example, cancer, given CYP2A6’s role in metabolizing various drugs, including the chemotherapeutic agents letrozole and tegafur [23, 24].

## Materials and methods

### Study samples

#### UK Biobank

UK Biobank (UKB) is a population-based prospective study on genetic and non-genetic determinants of diseases of middle and old age. The cohort consists of over 500,000 participants from the UK, aged between 37–73 at recruitment (2006–2010) [25]. The resource comprises imputed genome-wide genotype data from all participants [26], along with a comprehensive range of phenotypic data. This includes data from clinical assessments, questionnaires, sample assays, and health record linkage—many of which are available from all of the participants (see Sudlow et al. [27]). Of note, the NMR is not available in the UKB data.

We had access to genetic data from 487,235 individuals after excluding withdrawals. We restricted our sample to individuals of self-reported White British ancestry, and performed further quality control measures, resulting in a final sample size of 343,662 individuals. During the quality control process, we ensured the genetic sex corresponded to the reported sex, and that there were no instances of sex aneuploidy. We also verified that our subset did not include any samples that were outliers with respect to genotype heterozygosity or missingness. Furthemore, we only retained one randomly chosen individual from each pair of third-degree or closer relatives.

#### FINRISK

The National FINRISK Study consists of cross-sectional population-based data on chronic non-communicable diseases in Finland [28]. Data collection was conducted every five years between 1972–2012. Our previous GWAS of the NMR (n = 5,185 current smokers from 5 cohorts) included 1,405 current smokers (Cot *≥* 10ng/ml) from the 2007 and 2012 FINRISK data collections [15]. These two cross-sectional studies included independent samples from 25–74 year old Finns (see Buchwald et al. [15], Table S1). Cot and 3HC concentrations were acquired from blood plasma samples using gas chromatography-mass spectrometry. Self-reported variables on smoking behaviour, including the number of factory and self-rolled cigarettes smoked per day (CPD), were obtained from surveys. Genome-wide genotype data were imputed using a Finnish reference panel (see Buchwald et al. [15], Table S2).

#### Young Finns Study

The Young Finns Study (YFS) is a prospective population-based study of cardiovascular risk factors from childhood to adulthood. The initial cross-sectional sample from 1980 was selected so as to be representative of Finnish children aged 3, 6, 9, 12, 15 and 18. The individuals have been followed up at regular intervals (see Raitakari et al. [29]).

Our previous GWAS of the NMR included 714 current smokers from the YFS, aged 15–45 (see Buchwald et al. [15], Table S1). For each individual we had chosen the time point for which the NMR, sex, age and BMI were available, Cot was *≥* 10ng/ml, and for which we had the least amount of missing values concerning the other GWAS variables (CPD, Pack years, alcohol use). Whenever there were ties, the most recent time point had been chosen. Cot and 3HC concentrations were acquired from blood plasma samples using liquid chromatography-tandem mass spectrometry, and self-reported smoking behaviour variables including CPD were obtained from surveys. Genome-wide genotype data was imputed using the Haplotype Reference Consortium reference panel (see Buchwald et al. [15], Table S2).

### Measures

#### The genetic score for the nicotine metabolite ratio

Using the UKB data we created a genetic score (GS) for the NMR. We created it so that higher values reflect faster nicotine metabolism, and thus, also refer to it in this paper as the genetic score for faster nicotine metabolism. The GS served as the independent variable (exposure) in our PheWAS. We constructed the GS as the weighted sum of the 10 putative causal SNPs highlighted by our FINEMAP analyses (see below). These SNPs together explained 33.8 % of the variance in the NMR. Prior to running the PheWAS, we standardized the GS (zGS) by subtracting the mean and dividing by the standard deviation for a simpler interpretation of the results.

#### Outcome variables

We used the PheWAS software package PHESANT [21] to preprocess the phenotype data. Of the 5,559 UKB phenotype fields available to us, we included 4,546 in our PheWAS. The excluded phenotype fields were either not listed in the variable information file of PHESANT (252 dropped), were categorized as auxiliary variables in the UKB data (STRATA=Auxiliary) (further 398 dropped) or based on the grouping done by Gibson et al. in their insomnia PheWAS [30] (further 141 dropped), or were marked for exclusion in the PHESANT variable information file (further 60 dropped).

PHESANT uses a rule-based system for preprocessing and deciding which association test to use for each variable. For full details, see [21]. For example, when a phenotype has been measured at multiple time points, PHESANT automatically considers only the first occurrence. For phenotypes measured multiple times at the first occurrence to improve accuracy, PHESANT uses the mean value of those measurements. For multiple choice questions where individuals have ticked all relevant options, PHESANT forms multiple binary variables. For continuous variables, PHESANT inverse rank transforms them to normality (unless, for example, there is a notable portion of some particular value, in which case an ordered categorical variable is created). Out of the 4,546 phenotype fields, PHESANT created 21,094 outcome variables in total, meaning that our initial PheWAS consisted of 21,094 regression analyses. Each outcome variable served as the dependent variable in its respective regression model.

#### Covariates

We adjusted our PheWAS for sex and age to increase statistical power and to adjust for potential confounding. We also adjusted for the first ten principal components of genetic structure (1–10 PCs) to control for confounding due to population stratification. We did not include any additional covariates to avoid inducing collider bias.

#### Ever-Never status

We wanted to perform the PheWAS separately for ever and never smokers to distinguish whether the associations were independent of nicotine metabolism or indicative of a possible causal pathway through nicotine metabolism (Fig 1), an approach described by Millard et al. [22]. To reduce noise, we excluded experimenters and occasional smokers from our Ever and Never subsets. We created a new Ever-Never variable based on two existing variables:

1. Current tobacco smoking (”Do you smoke tobacco now?”, UKB field 1239)
2. Past tobacco smoking (”In the past, how often have you smoked tobacco?”, UKB field 1249)

**Fig 1.**
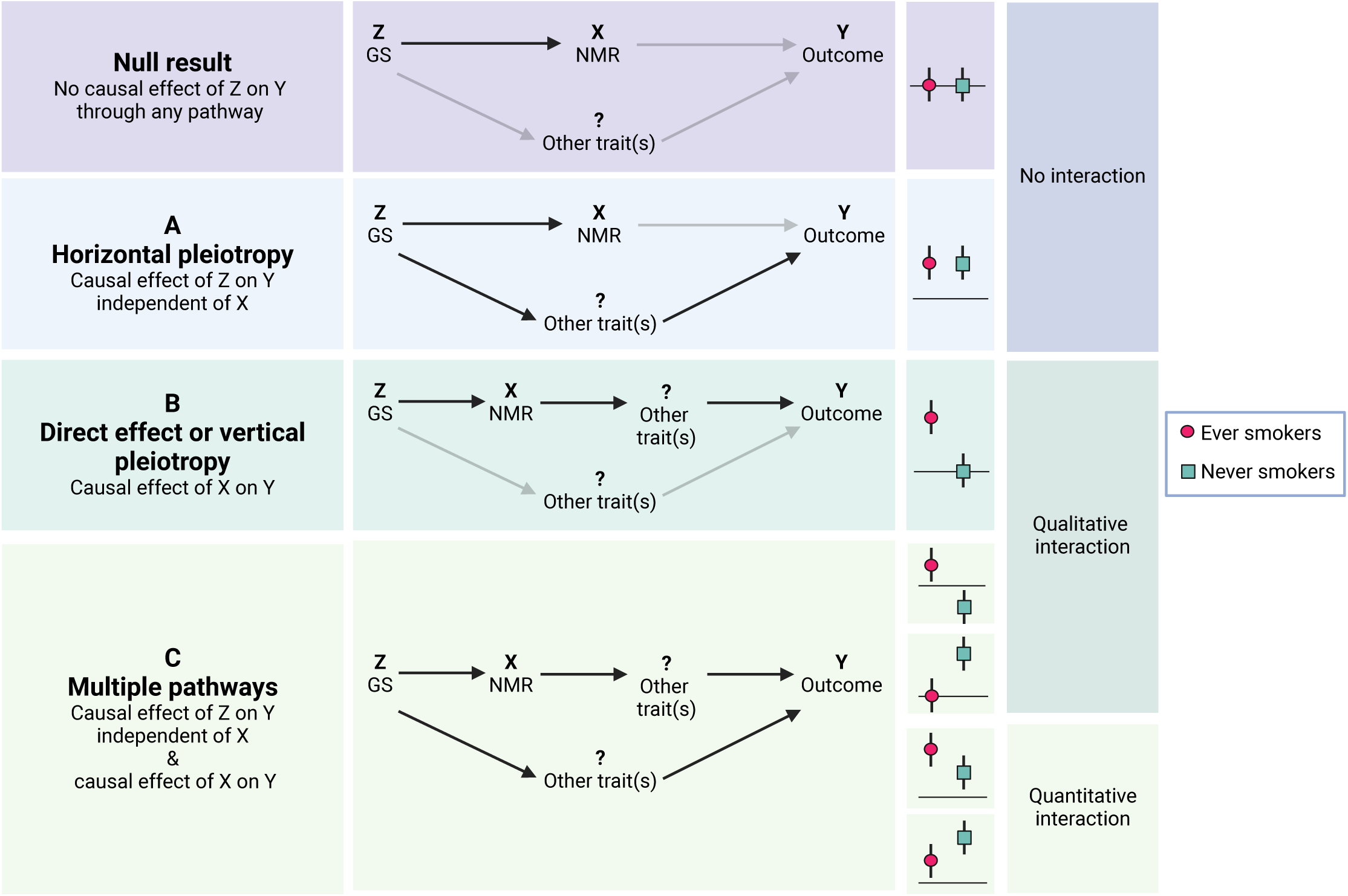
Interpretations of the possible GxE MR results. The figure illustrates how the different types of results from GxE MR can be interpreted in our case, where the exposure, X, is the NMR, and the genetic instrumental variable, Z, is the genetic score for the NMR. The ever and never results can be categorized in to three types: 1) no interaction, i.e. the effect of Z on outcome Y is not affected by smoking status, 2) qualitative interaction, i.e. the effect of Z on Y is apparent only in one group or the effects are in opposing directions between the groups, and 3) quantitative interaction, i.e. the effects are consistent between the groups but of different magnitude. There are three different interpretations for the associations: A) if an effect of the same magnitude is apparent in both groups, there is evidence for a causal effect of Z on Y through a pathway excluding X (this is known as horizontal pleiotropy), B) if an association is only seen among ever smokers, there is evidence for a causal effect of Z on Y through X, and for X being causally associated with Y, either directly or through another trait/other traits such as amount smoked (the latter is known as vertical pleiotropy), and C) if the ever and never subsets show opposing effects, effects of different magnitude, or there is only an effect among never smokers, there is evidence for a causal effect of Z both through the NMR (either directly or through vertical pleiotropy) and through another pathway (horizontal pleiotropy). The figure has been modified from Millard et al. Fig 1 [22] and the figures at: https://mr-dictionary.mrcieu.ac.uk/term/vertical-pleiotropy/. The figure was created with BioRender.com.

The second question had been asked from all except for those who indicated they currently smoke on “most or all days” in response to the first question. We classified all who answered “most or all days” to either of the two questions as ever smokers. Those who answered “no” to the first question and “never” to the second question, were classified as never smokers. All others were assigned a missing value in our Ever–Never variable, and excluded from the Ever and Never subsets. This included individuals who answered “Occasionally” or “Prefer not to answer” to either question, as well as those who responded with “Tried once or twice” to the second question.

Out of our full sample (n = 343,662), we classified 135,890 as never smokers and 110,348 as ever smokers. Of those who dropped out, 99 % had answered “Occasionally” or “Tried once or twice”. This group had marginally higher GS values as compared to the Never group (Table S1 & Fig. S2). The GS distribution of the Ever group did not differ from the Never group (Table S1 & Fig. S2).

### Statistical analyses

Fig. S1 illustrates the main analyses and follow-up analyses performed in this study.

#### Forming the GS

We previously performed a GWAS meta-analysis of the NMR in European ancestry current smokers (n = 5,185) [15]. Our main GWAS model included sex, age, BMI, alcohol use (g/week) and birth year as covariates. We identified two NMR loci, one on chromosome 4 and one on chromosome 19. By fine-mapping these loci using FINEMAP [31] and the datasets YFS and FINRISK (n = 2,119), we found there to be only a single putative causal SNP on chromosome 4, explaining 1.7 % of the variance in the NMR, but 13 putative causal SNPs on chromosome 19 explaining 36.7 % of the variance in the NMR.

In the current study, we wanted to use the SNPs and weights obtained by the earlier FINEMAP analyses to construct the GS for the NMR in the UKB data set. Unfortunately, two of the 13 SNPs from chromosome 19 did not pass our quality control in the UKB dataset (both had a Hardy-Weinberg Equilibrium *p <* 10*^−^*^6^). Excluding these two SNPs would have dropped the variance explained in the NMR by the chromosome 19 SNPs from 36.7 % to 23.7 %. Therefore, to gain maximal power, we reran the FINEMAP analyses for the chromosome 19 locus as described previously (see Buchwald et al. [15]) using only SNPs that were available and passed quality control in UKB (See Supplementary information). The new FINEMAP analysis of the chromosome 19 locus resulted in nine SNPs in the top configuration of putative causal SNPs, explaining 32.1 % of the variance in the NMR (Table S2). All the aforementioned estimates of the variance of the NMR explained, are the configuration-specific heritability estimates obtained by FINEMAP.

We then calculated the GS as a weighted sum of these nine chromosome 19 SNPs and a single chromosome 4 SNP, resulting in a GS explaining 33.8 % of the variance in the NMR (Table 1). For both loci, we used the largest possible sample sizes to estimate the effect sizes with the highest possible accuracy. For chromosome 19, we used the joint effect size estimates from FINEMAP as the weights in order to adjust for correlation among these SNPs. As only the datasets YFS and FINRISK were available for fine-mapping, the sample size was n = 2,119. For chromosome 4, as there was only one putative causal SNP, and thus no need to adjust for any correlated SNPs, the effect estimate of this single chromosome 4 SNP equals to its marginal effect in the original GWAS (n = 5,185). Of note, FINEMAP was not able to distinguish which of the three top SNPs in the chromosome 4 locus was most likely to be the causal one [15]. All three SNPs had the same p-value and were perfectly correlated. For the GS of the NMR, we chose to use the SNP located in the middle based on base pair position.

**Table 1.** The ten SNPs and their weights used for calculating the GS for the NMR.

| SNP | CHR | BP | EA/NEA | MINOR | MAF<sub>UKB</sub> | WEIGHT (MAF) |
| --- | --- | --- | --- | --- | --- | --- |
| RS36103218 | 4 | 69359253 | T/C | C | 0.3422 | -0.175 (MAF<sub>META</sub> = 0.43) |
| RS189621498 | 19 | 41288136 | A/G | A | 0.0003 | 0.6127 (MAF<sub>FY</sub> = 0.03) |
| RS74719953 | 19 | 41335799 | T/C | T | 0.0652 | -0.3581 (MAF<sub>FY</sub> = 0.09) |
| RS34945948 | 19 | 41340842 | G/A | G | 0.1474 | 1.0276 (MAF<sub>FY</sub> = 0.14) |
| RS12985907 | 19 | 41343544 | A/G | A | 0.2307 | -0.8003 (MAF<sub>FY</sub> = 0.28) |
| RS1801272 | 19 | 41354533 | T/A | T | 0.0271 | -1.0103 (MAF<sub>FY</sub> = 0.02) |
| RS7250713 | 19 | 41355195 | C/G | G | 0.4386 | 0.3251 (MAF<sub>FY</sub> = 0.37) |
| RS7248187 | 19 | 41437426 | C/G | G | 0.3023 | 0.3187 (MAF<sub>FY</sub> = 0.24) |
| RS116382863 | 19 | 41534881 | T/C | T | 0.1242 | 0.2212 (MAF<sub>FY</sub> = 0.13) |
| RS11466310 | 19 | 41861858 | T/C | T | 0.0244 | -0.5414 (MAF<sub>FY</sub> = 0.02) |

To assess the performance and reliability of our GS, we plotted it against the NMR top SNP (rs56113850) in the UKB data. In our previous study, the top SNP alone explained 23 % of the variance in the NMR (see Buchwald et al [15], Table S5). Among current smokers, we also plotted cigarettes smoked per day (CPD) against our GS as CPD has been shown to associate positively with the NMR [8, 9]. Additionally, we tested the association of the GS with CPD with a linear model. We did this by first regressing out sex, age and the first ten principal components of genetic structure, and then inverse normalizing CPD. We used the whole data but also split the data into two: lower and higher ends of the GS. We used 0 as the cut-off point based on the bend apparent in the loess curve of the scatter plot (Fig 2) and to obtain roughly the same sized groups as the mean was close to the median (Table 2). In order to have a reference point, we used our Finnish data to create plots of CPD against the NMR, as well as Cot+3HC, a biomarker for nicotine intake, against the NMR.

**Fig 2.**
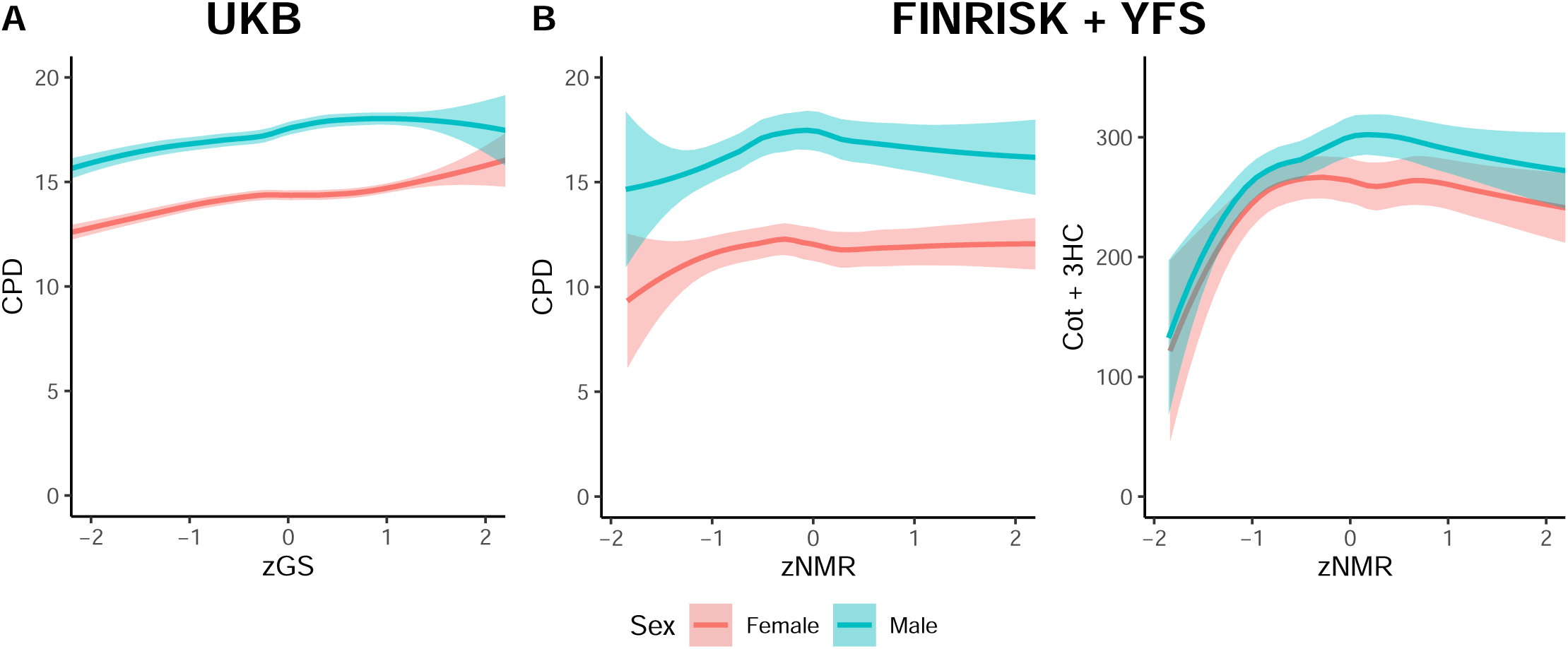
**A** Loess curve depicting the association between the standardized genetic score (zGS) for the Nicotine Metabolite Ratio (NMR) and cigarettes smoked per day (CPD) in UK Biobank (UKB). **B** Loess curves depicting the associations between the standardized NMR (zNMR) and CPD, as well as zNMR and Cotinine + 3-Hydroxycotinine (Cot + 3HC), a biomarker for nicotine intake, using the combined data from FINRISK and Young Finns Study (YFS). All plots are for current smokers. The x-axes have been restricted to show only data points within two standard deviations from the mean to highlight the main trends. Full data are shown in Fig. S4. The standardized variables (zGS and zNMR) were calculated by subtracting the mean and dividing by the standard deviation.

**Table 2.** Descriptive statistics for age, sex, the GS for faster nicotine metabolism, and various smoking variables in the UKB data.

|  | All<br>n = 343,662 | Female<br>n = 184,565 (53.7%) | Male<br>n = 159,097 (46.3%) |
| --- | --- | --- | --- |
|  | mean (sd), median [min, max] | mean (sd), median [min, max] | mean (sd), median [min, max] |
| Age | 56.9 (8), 58 [39, 72] | 56.7 (7.9), 58 [40, 71] | 57.1 (8.1), 59 [39, 72] |
| GS | 1.2 (0.6), 1.2 [-1.7, 2.9] | 1.2 (0.6), 1.2 [-1.7, 2.9] | 1.2 (0.6), 1.2 [-1.7, 2.8] |
| zGS | 0 (1), 0.1 [-5.2, 3.1] | 0 (1), 0.1 [-5.2, 3.1] | 0 (1), 0.1 [-5.2, 2.9] |
|  | n (%) | n (%) | n (%) |
| **Smoking status** |  |  |  |
| Never | 135,890 (39.7) | 81,037 (44.1) | 54,853 (34.6) |
| Experimenter | 96,272 (28.1) | 52,700 (28.6) | 43,572 (27.5) |
| Ever | 110,348 (32.2) | 50,217 (27.3) | 60,131 (38.0) |
| **Former-Current (Ever subset)** |  |  |  |
| Former | 81,179 (73.6) | 36,490 (72.7) | 44,689 (74.3) |
| Current | 29,141 (26.4) | 13,711 (27.3) | 15,430 (25.7) |
| **Wants to quit (Current subset)** |  |  |  |
| Yes | 17,876 (70.3) | 8,759 (72.4) | 9,117 (68.4) |
| No | 7,543 (29.7) | 3,336 (27.6) | 4,207 (31.6) |
|  | mean (sd), median [min, max] | mean (sd), median [min, max] | mean (sd), median [min, max] |
| **Age started smoking on most days** |  |  |  |
| Current (n = 25,522) | 17.7 (5.8), 16 [5, 69] | 18.1 (5.8), 17 [5, 69] | 17.4 (5.7), 16 [5, 65] |
| Former (n = 80,788) | 17.2 (3.6), 17 [5, 63] | 17.7 (3.7), 17 [5, 60] | 16.8 (3.4), 16 [5, 63] |
| **Cigarettes smoked per day** |  |  |  |
| Current (n = 23,682) | 15.7 (8.4), 15 [1, 140] | 14.2 (7.3), 15 [1, 120] | 17.4 (9.2), 15 [1, 140] |
| Former (n = 76,865) | 19.3 (10.5), 20 [1, 140] | 16.8 (8.4), 15 [1, 100] | 21.5 (11.6), 20 [1, 140] |
| **Number of unsuccessful stop-smoking attempts** |  |  |  |
| Current (n = 2,527) | 4.1 (10.1), 3 [0, 200] | 4 (10.7), 3 [0, 200] | 4.1 (9.7), 3 [0, 200] |
| Former (n = 74,108) | 2.9 (7.1), 2 [0, 200] | 2.4 (5.3), 2 [0, 200] | 3.2 (8.3), 2 [0, 200] |
| **Age stopped smoking on most days** |  |  |  |
| Current (n = 3,218) | 47.3 (11.6), 48 [12, 70] | 47.4 (11.5), 48 [13, 70] | 47.1 (11.6), 48 [12, 70] |
| Former (n = 80,859) | 39.5 (11.6), 39 [9, 69] | 39.2 (11.5), 38 [12, 69] | 39.7 (11.6), 39 [9, 69] |

*GS*, genetic score for the nicotine metabolite ratio; *zGS*, standardized GS; *Ever*, individuals indicating smoking on most days at present (UKB Field 1239) or in the past (UKB Field 1249); *Never*, individuals responding negatively to current (UKB Field 1239) and past (UKB Field 1249) tobacco smoking; *Experimenters*, individuals indicating occasional or minimal past (UKB Field 1249) or present (UKB Field 1239) tobacco use; *Former-Current*, subsets of ever smokers differentiating those who had smoked in the past and those who smoke presently based on UKB Field 20116; *Wants to quit*, derived from the variable “Wants to stop smoking” (UKB Field 3496) with responses grouped into “Yes” (including “Yes, definitely” and “Yes, probably”) and “No” (including “No, probably not” and “No, definitely not”); *Age started smoking on most days*, UKB Field 2867 for former smokers, UKB Field 3436 for current smokers; *Cigarettes smoked per day*, UKB Field 3456 for current smokers, UKB Field 2887 for former smokers; *Number of unsuccessful stop-smoking attempts*, all individuals who had indicated that in the past they smoked tobacco on most or all days and that during the time they smoked they stopped for more than 6 months were asked “How many times did you try to give up smoking before you were successful?” (UKB Field 2926); *Age stopped smoking on most days*, UKB Field 2897.

For all chromosome 19 SNPs the weight has been obtained from their joint model using FINEMAP in the Finnish data. For the chromosome 4 SNP the weight is the effect size obtained from our previous GWAS [15]. *SNP*, single-nucleotide polymorphism; *CHR*, chromosome; *BP*, base pair position in GRCh37 coordinates; *EA/NEA*, the effect allele/ the non-effect allele; *MINOR*, the less common allele; *MAF*, minor allele frequency; *WEIGHT*, weight used to calculate the GS (reported for the effect allele); *UKB*, UK Biobank; *META*, data used in our previous GWAS meta-analysis (n = 5,185) from which the weight for the chr 4 SNP is; *FY*, FINRISK and YFS data (n = 2,119) used to obtain the weights for the chr 19 SNPs.

#### PheWAS

We used the GxE MR-pheWAS approach [22], which allows for hypothesis-free testing of the causal effects of the exposure (in our case, the NMR), while simultaneously examining the presence of association pathways not involving the exposure (horizontal pleiotropy) (See Supplementary Information). The idea is to divide the data in to groups with different levels of the exposure, in our case, ever and never smokers. We assume that the effect of nicotine metabolism only occurs in people who are actually using nicotine, in other words our ever smokers subset. The approach permits us to distinguish whether the PheWAS associations reflect a causal pathway through a) the NMR, either directly or through other traits such as amount smoked (effect only seen in ever smokers), b) some other pathway not including the NMR (same effect seen also in never smokers), or c) both (effect only seen in never smokers, or there are quantitative or directional differences in the effect sizes between ever and never smokers) (see Fig 1).

We began by running PHESANT (version 1.1) on the full sample using the ‘save’ option to save the variables processed and derived by PHESANT. Out of the 4,546 UKB phenotype fields we had to begin with, PHESANT created a total of 21,094 outcome variables. We then performed the actual PheWAS analyses in two stages.

#### Exploratory PheWAS

In the first stage, we used the 21,094 PHESANT derived outcome variables to run an exploratory PheWAS for the entire sample (All), as well as separately for those who had ever smoked daily (Ever) and those who had never smoked (Never). We adjusted for age, sex and 1-10 PCs. For each PheWAS (All/Ever/Never), we used the 5 % false discovery rate (FDR) level to define statistical significance at the phenome-wide level. We used the Benjamini-Hochberg method to obtain the cut-off points for the 5 % FDR, i.e. the thresholds for phenome-wide significance (TPWSs).

Within each PheWAS, we ranked the results based on ascending p-values. Next, for each outcome variable, we calculated the critical value *c_i_* = 0.05 *× i/n*, where *i* is the rank and *n* the total number of tests performed in that PheWAS. The TPWS, was then defined as the *c_i_* corresponding to the outcome with the highest rank to satisfy the condition *p_i_ ≤ c_i_* [32]. We used these TPWSs also in the second stage.

#### Final PheWAS

In the second stage of our analyses, we wanted to confirm our findings.

We reran the regression analyses for all the outcomes with phenome-wide significant (PWS) associations in any of the three exploratory PheWASs described above (All/Ever/Never). First, we manually checked the coding and the distributions of these highlighted variables, as well as the appropriateness of the models used by PHESANT. Where appropriate, we split the variable into multiple variables, adjusted the coding or used a different regression model. We did these changes to obtain results that are more readily interpretable. Additionally, we reran the highlighted linear regression analyses by regressing the covariates (age, sex, 1-10 PCs) out before inverse rank transforming the outcome variables to normality. We did this because many of these continuous outcomes, such as waist circumference, are known to be normally distributed around different values depending on sex (one of our covariates). For the linear models, we didn’t adjust for covariates anymore in the actual regression analyses. Otherwise, we used the same protocol as PHESANT for our final analyses of the All, Ever and Never data sets. A detailed description of the second stage outcomes that we derived, recoded or analysed using a different model from the initial exploratory PheWASs can be found in Table S3. For each data set we used the corresponding TPWS obtained previously to determine statistical significance at the phenome-wide level.

#### Differences between the ever and never smokers

We wanted to assess whether there were any outcome variables for which there was a statistically significant difference between the effect sizes for the Ever and Never subsets. We did the Ever versus Never analyses in R by deriving p-values for the differences using the function pchisq(x, df=1, lower=FALSE), where

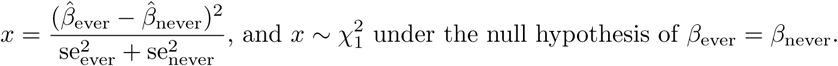

First, we did this for our initial PheWAS results. Of all the 21,094 PHESANT derived outcomes altogether 15,212 had been analysed for both the Ever and Never subsets using the same regression model. Once again, we used the 5 % FDR level to determine phenome-wide significance, and obtained the TPWS using the Benjamini-Hochberg method. We repeated the Ever versus Never analyses for our outcomes from the second stage of our PheWAS analyses, using this same TPWS to define statistical significance.

### Follow-up analyses

#### Current versus Former analysis

We followed-up our Ever versus Never analyses by taking all the outcomes with a PWS difference in their effect sizes, and repeated the analysis for current versus former smokers. We did this to obtain a better understanding of the possible causal role of smoking in the association pathway. We constructed the current and former smoker subgroups from our Ever subset, and repeated the second stage PheWAS regression analyses for the chosen outcomes in these two groups.

#### Follow-up of the GS-Cessation association

Our PheWAS results suggested a positive association between the GS for faster nicotine metabolism and smoking cessation. To investigate further what could be driving the positive association, we examined the variable “Why did you stop smoking? (You can select more than one answer): Illness or ill health / Doctor’s advice / Health precaution / Financial reasons” (UKB Field 6157), and the variable “Number of unsuccessful stop-smoking attempts” (UKB Field 2926) which captures the difficulty of quitting.

As faster nicotine metabolism is thought to associate with a greater amount of cigarettes smoked and more difficulties in quitting smoking successfully (reviewed in [10]), we began by rerunning the logistic regression model for cessation, including CPD as an additional covariate to adjust for possible confounding. In other words, among ever smokers, we ran a logistic regression, where cessation was the dependent variable, and the zGS, age, sex, 1–10 PCs, and CPD were the predictor variables. Smoking more will increase the risk of severe illnesses and the need for treatments such as surgery that require abstinence. Thus, we also ran this model within a subset of the ever smokers from whom we had excluded those who listed illness and/or doctor’s advice as the reason for stopping smoking.

Reasons for stopping smoking were available for those ever smokers who had at least once managed to stop for over six months. We divided the Ever subset into three approximately equal sized groups based on the GS tertiles, and named these groups as slow, medium and fast metabolizers. We then compared, one reason at a time, whether the proportions differed between the slow and fast groups. We used the 2-sample test for equality of proportions using the prop.test function in R.

To assess the importance of the different reasons, we reran the cessation models (with and without CPD included as a predictor) among those ever smokers who had at least once managed to stop for over six months. Among this subset, we added all four reasons to the models. We wanted to see how the reasons predicted former status at the time of the questionnaire.

Then, among our former smokers, we ran a negative binomial regression model with “Number of unsuccessful stop-smoking attempts” as the dependent variable and each of the four reasons, the zGS, age, sex, and 1–10 PCs as the predictor variables. We also ran the model including CPD as a confounder.

#### MRBase and FinnGen PheWASs using the top SNP for the NMR

For comparison, we ran PheWASs of the top SNP for the NMR (rs56113850) using GWAS summary data from FinnGen (Data freeze 9, from April 2022) and MRBase (Database version 0.3.0, from 25 Oct 2020). We used the open access web interfaces. The FinnGen Data Freeze 9 has a total sample size of 377,277 and consists of 2,272 disease endpoints from Finnish biobank participants [33, 34]. The GWAS results from FinnGen were adjusted for sex, age, PC1-10, whether using FINNGEN1 or 2 chip, and legacy genotyping batch [33]. The MRBase comprises GWAS summary data from numerous consortia, including UKB and FinnGen [35, 36]. From MRBase, we got results for 39,105 outcomes. This group of outcomes included expression quantitative trait loci (eQTLs) as well. Unfortunately, we do not know what the GWAS results from MRBase were adjusted for as the results come from multiple different cohorts and studies using different protocols.

In both cases, we defined phenome-wide significance as before, and created tables of the PWS results, i.e. those reaching statistical significance at the 5 % FDR level using the Benjamini-Hochberg method. We used the library biomaRt in R to annotate the eQTLs that were highlighted in the MRBase PheWAS.

#### Assessing sex and ancestry differences

We scrutinized our results by exploring possible differences between sexes and among ancestries. First, we repeated the final PheWAS analyses exactly as before, this time stratifying by sex. We then assessed whether the effect sizes differed between the sexes by using the same formula we used for the ever-never comparisons. Second, to assess to which extent our results can be generalized to other ethnic groups, we used the mentioned top SNP for the NMR, rs56113850-allele C (versus T), as a proxy for genetically determined faster nicotine metabolism. This SNP has been shown to replicate in other ancestries, and to be an important predictor for the NMR across ancestries [37–39]. We reran all the 33 continuous phenotypes highlighted as having a PWS association in our final PheWAS, this time stratifying by ancestry and using the top SNP instead of the GS. We focused on the continuous traits, since the sample sizes in the ancestries other than White British were too small for disease studies (Table S11). To maintain reasonable sample sizes, we did not separate ever and never smokers in the ancestry-specific analyses. To create the ancestry groups that we then compared to our White British sample, we used the Top level groupings listed for the UKB field 21000 on showcase [40].

## Results

### Descriptive statistics

For female participants, the smoking status (Ever/Experimenter/Never) category Never was the largest (44 %), whilst for males the Ever category was the largest (38 %) (Table 2). Among both sexes, over 70 % of the ever smokers were former smokers, and roughly 70 % of the current smokers wanted to stop smoking. The distribution of the GS for the NMR was nearly identical for both sexes (Table 2). When running a linear model of the standardized GS by age, the age variable was not statistically significant (*p* = 0.909).

The correlation between the GS and the top SNP (rs56113850) was 0.73 (Fig. S3). Each standard deviation increase in the GS was associated with an increase of 0.6 cigarettes smoked per day (CPD), after adjusting for age, sex and 1–10 PCs (Table S4). The association seemed to be stronger at the lower end of the GS than at the higher end (Table S4, Fig 2a). This trend could also be seen in our Finnish sample when plotting CPD or Cot+3HC against the NMR (Fig 2b).

### Exploratory PheWAS

A total of 61 different outcomes reached phenome-wide significance across the initial All, Ever, and/or Never PheWASs (Fig. S5, Table S5). The exploratory full sample PheWAS resulted in 47 PWS associations (TPWS_All_: 0.05 *·* 47*/*21094 = 1.1*e −* 04) (Table S5a). For the Ever subset there were 29 PWS associations (TPWS_Ever_: 0.05 *·* 29*/*16648 = 8.7*e −* 05) (Table S5b), and for the Never subset, only two associations were PWS (TPWS_Never_: 0.05 *·* 2*/*16103 = 6.2*e −* 06) (Table S5c).

### Final PheWAS

The set of 61 outcome variables highlighted in the first stage of our PheWAS analyses was then taken to the second stage. Some of these variables were split into multiple variables to tease out which aspect of the phenotype was driving the association (Table S3). For example, Smoking Status (Never/Previous/Current) was split into Cessation (former vs current smoker) and Initiation (ever vs never smoker), out of which only Cessation showed a PWS association when we reran the analyses (Fig 3).

**Fig 3.**
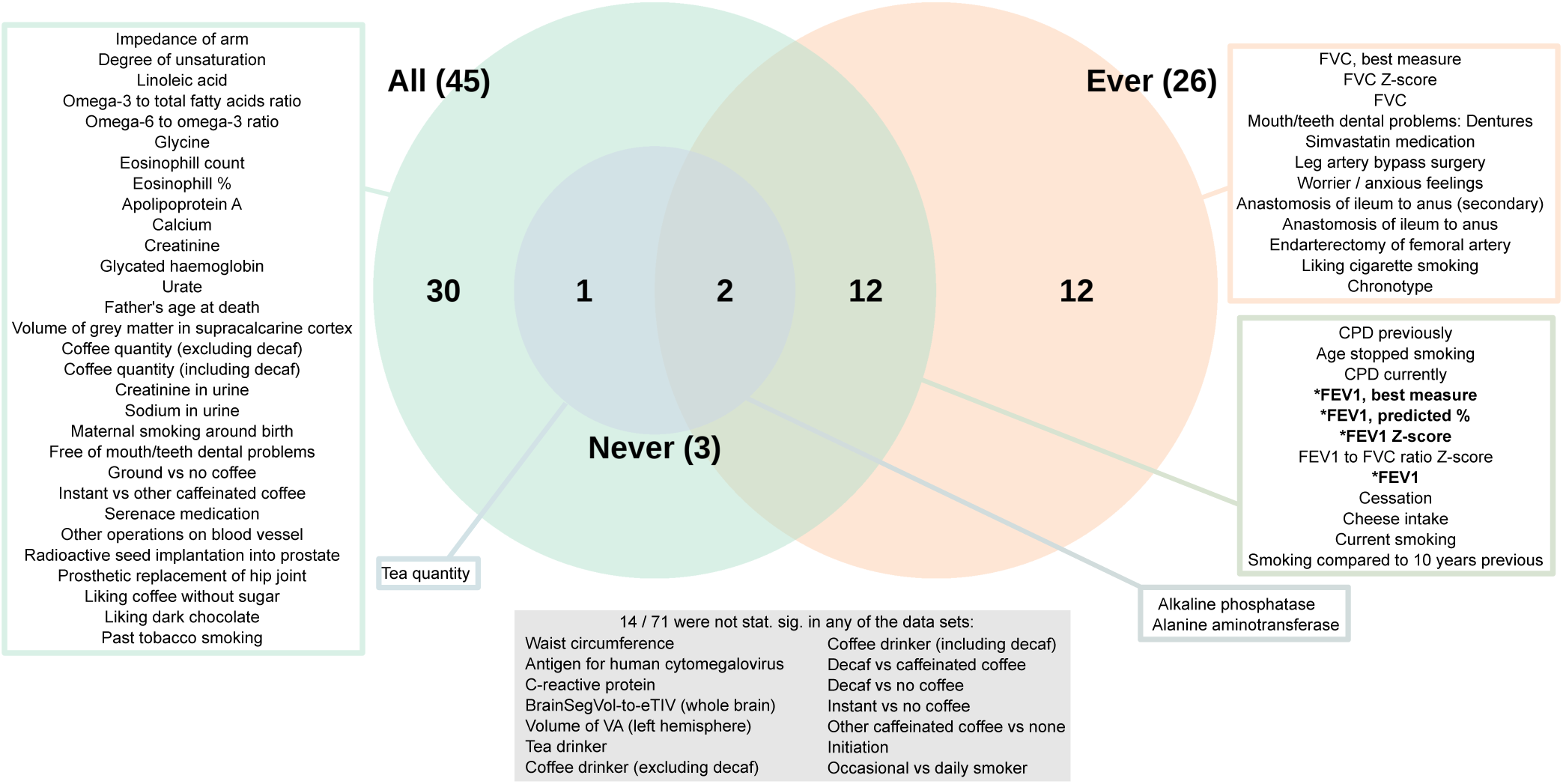
Venn diagram of the results of the final 71 variables. Out of these variables, 57 had a phenome-wide significant (PWS) association in at least one of the data sets (All / Ever /Never). Those with a PWS difference in their effect sizes between the ever and never smokers have been indicated with a star and bolded text.

Our second stage of the PheWAS analyses included 71 outcome variables (Table S6). Of these, 57 showed a PWS association in at least one of the three data sets based on the data specific TPWSs (Fig 3, Fig 4). We had 45 PWS associations among the full sample, 26 among the Ever subset and three among the Never subset (Fig 3, Table S6b–d).

**Fig 4.**
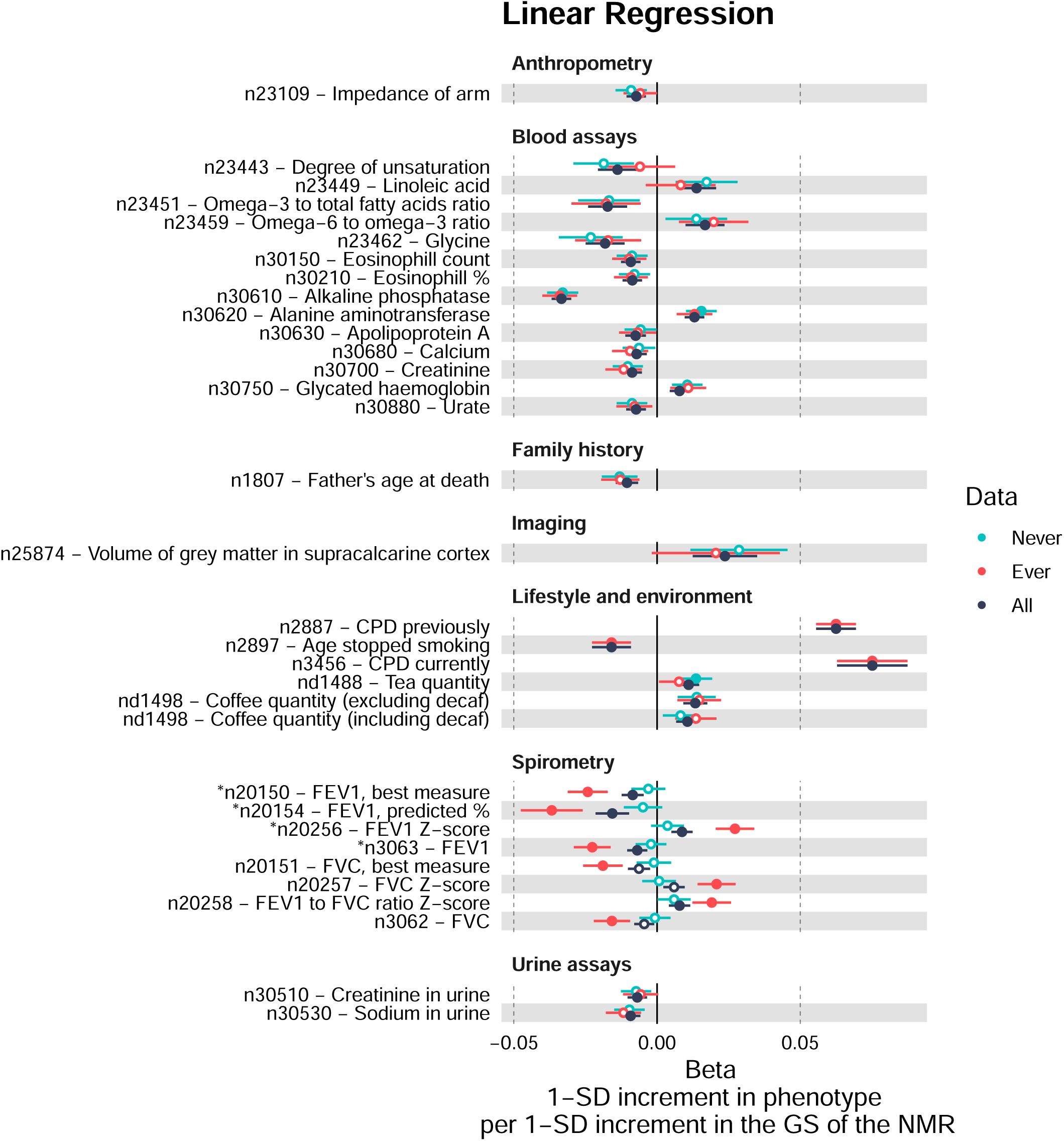
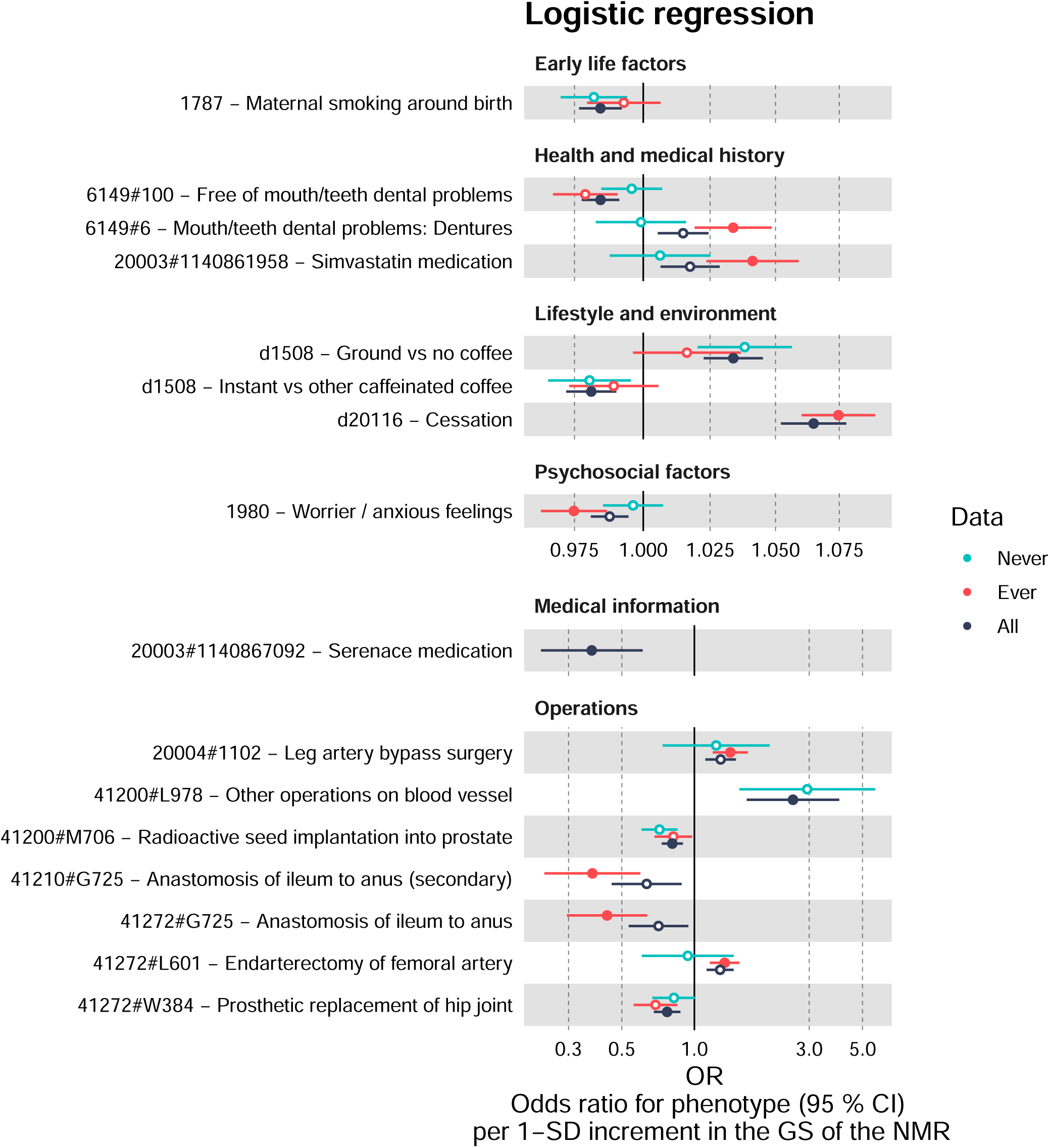
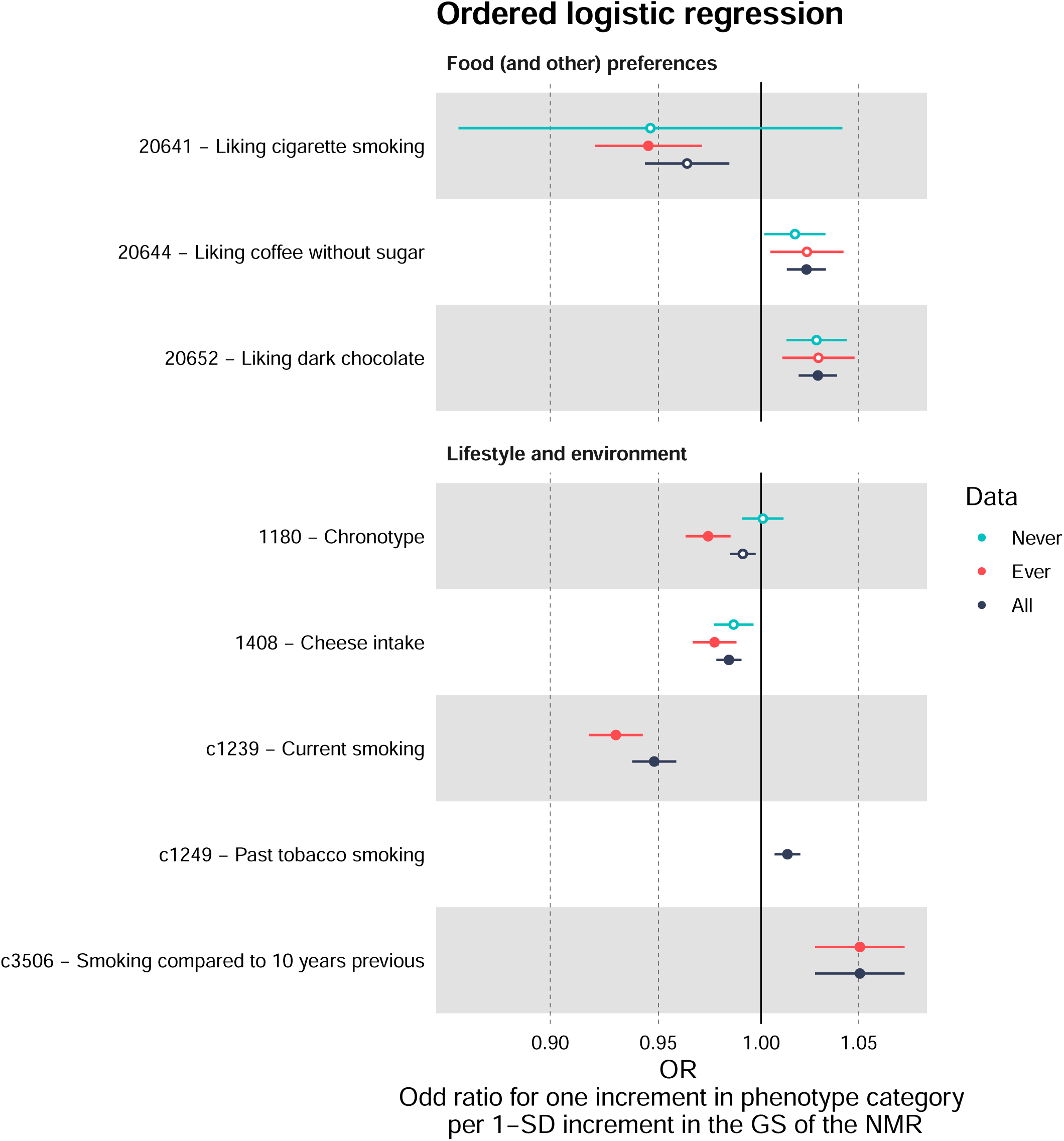
Results of the 71 variables from the final PheWAS analyses. The results of those variables (n = 57) that were phenome-wide significant (PWS) (solid circles) in at least one of the data sets (All / Ever / Never) are shown. *, there was a PWS difference between the effect sizes of the ever and never smokers; *n*, normalised after covariates had first been regressed out; *d*, derived from the original UKB phenotype; *c*, coding corrected to be more intuitive.

For 14 outcomes, we did not observe a PWS association with the GS for faster nicotine metabolism in any of the three groups (All, Ever or Never). This set of non-significant associations included five continuous variables that had reached the TPWS in at least one of the data sets in the exploratory PheWASs (when the covariates had not been adjusted for before normalizing the outcome). The rest of the non-significant results included the derived Smoking Initiation and Occasional vs Daily Smoker variables, as well as some of the derived tea and coffee variables.

### Ever versus Never analyses

In the first stage of our PheWAS, a total of 15,212 outcome variables had been analysed for both the Ever and Never subsets using the same regression model. Of these, only two variables showed a PWS difference in their effect sizes between the ever and never smokers (Table S5d). The TPWS was thus at 0.05 *·* 2*/*15212 = 6.6*e −* 06. Both of these were lung capacity variables and were included among the variables highlighted in the exploratory All and Ever PheWASs (Fig. S5). There was no association among the Never subset while among the Ever and All groups higher values of the GS predicted worse lung capacity.

Of the 71 outcome variables included in the second stage of our PheWAS analyses, 58 were available for our Ever versus Never analysis. Of these, four showed a PWS difference between ever and never smokers in their effect sizes (*p <* 6.6*e −* 06) (Table S6a, Fig 4). All four were lung capacity measures. For all four, there was no evidence of association among never smokers but a PWS association among ever smokers, suggesting a causal pathway through smoking. The results suggested that only among ever smokers higher values of the GS predicted worse lung capacity.

### Never smokers: associations with liver enzymes and tea quantity

Among never smokers, a higher GS for faster nicotine metabolism was associated with decreased alkaline phosphatase and increased alanine aminotransferase, both of which are liver enzymes (Table S6d). These associations were also seen among the Ever and All groups (Fig 3, Fig 4), and were PWS for all three groups already in our exploratory PheWASs (Fig. S5).

Among never smokers, there was also a PWS association with increased tea consumption. This was PWS in the All group too, but not among the ever smokers subset. Apart from these three outcomes, none of the other outcomes showed a PWS association in the never smokers subset. The direction and magnitude of association was nevertheless similar to those of the other groups for many variables such as for the other blood and urine assay variables and variables related to coffee (Fig 4).

### Ever smokers: associations with liver enzymes and smoking related variables

Among ever smokers, 26 of the final second stage outcome variables showed PWS associations (Fig 3, Table S6c). In addition to the liver enzymes, a higher GS for faster nicotine metabolism was associated with a greater number of cigarettes smoked per day, an increased likelihood of having quit smoking, decreased lung capacity, an increased likelihood of taking cholesterol medicine (Simvastatin), being more of a morning person than an evening person, quitting smoking at a younger age, an increased likelihood of having dentures, increased smoking compared to 10 years ago, decreased consumption of cheese, a decreased likelihood of being a worrier, decreased liking for cigarette smoking, an increased likelihood of leg artery operations, and a decreased likelihood of an anastomosis of ileum to anus.

The GS for faster nicotine metabolism was also associated with an increased likelihood of smoking less at the time of the interview (”Do you smoke tobacco now?: No / Only occasionally / Yes, on most or all days”, UKB field 1239). This negative association with current smoking was driven by the higher odds of quitting. There was no association for the derived Occasional vs Daily outcome, whereas our derived Cessation outcome demonstrated one of the strongest associations (Table S6c). Our ever smokers only contained individuals who smoked daily or had smoked daily in the past. Therefore, we had better statistical power for our Occasional vs Daily phenotype in our full sample. Nevertheless, we saw no association there either (*p* = 0.90, Table S6b).

### Full sample: strongest associations with liver enzymes, smoking related variables, coffee, and tea

Among the full sample there were 45 outcome variables that showed a PWS association with the GS for the NMR (Fig 3, Table S6b). The strongest associations were observed for the liver enzymes, CPD, cessation, and coffee and tea consumption. The GS was also associated with 16 biomarkers, often used to assess overall health. For example, the GS was associated with outcomes related to fatty acids, blood sugar, kidney and liver health, impedance of arm, calcium, and white blood cells. The Never and Ever associations for these biomarkers were in line with each other but neither reached phenome-wide significance, except for the liver enzymes mentioned earlier (Fig 4).

Some additional outcome variables, that had not reached phenome-wide significance in the smaller subsets, were highlighted in the full sample. For example, among the full set, the GS for faster nicotine metabolism was associated with an increased liking for dark chocolate and coffee without sugar, decreased father’s age at death, decreased maternal smoking around birth, increased volumes of grey matter in the supracalcarine cortex, and a decreased likelihood of using haloperidol (Serenace), an anti-psychotic medication. Some outcomes were only analysed in some groups due to a lack of cases, and the use of haloperidol was one of these (only analysed in the full sample). The outcome Past tobacco smoking (Never/Tried once or twice/Occasionally/Most days or daily) was only analysed in the All group as it required answers from both ever and never smokers. The GS for faster nicotine metabolism was associated with an increased likelihood of being in a higher past tobacco smoking category.

### Follow-up analyses

#### Comparison of current and former smokers supports the causal role of smoking on worse lung functioning

Our Ever versus Never analyses highlighted four lung capacity measures, suggesting a causal pathway through nicotine metabolism, as the association was only seen in the ever smokers. We followed-up these results by rerunning the analyses for the current and former smoker subsets. The effect sizes for all four lung capacity measures were smaller among former smokers compared to current smokers, though they too remained PWS (*p <* TPWS_Ever_ = 8.7*e −* 05) (Fig 5, Table S6e).

**Fig 5.**
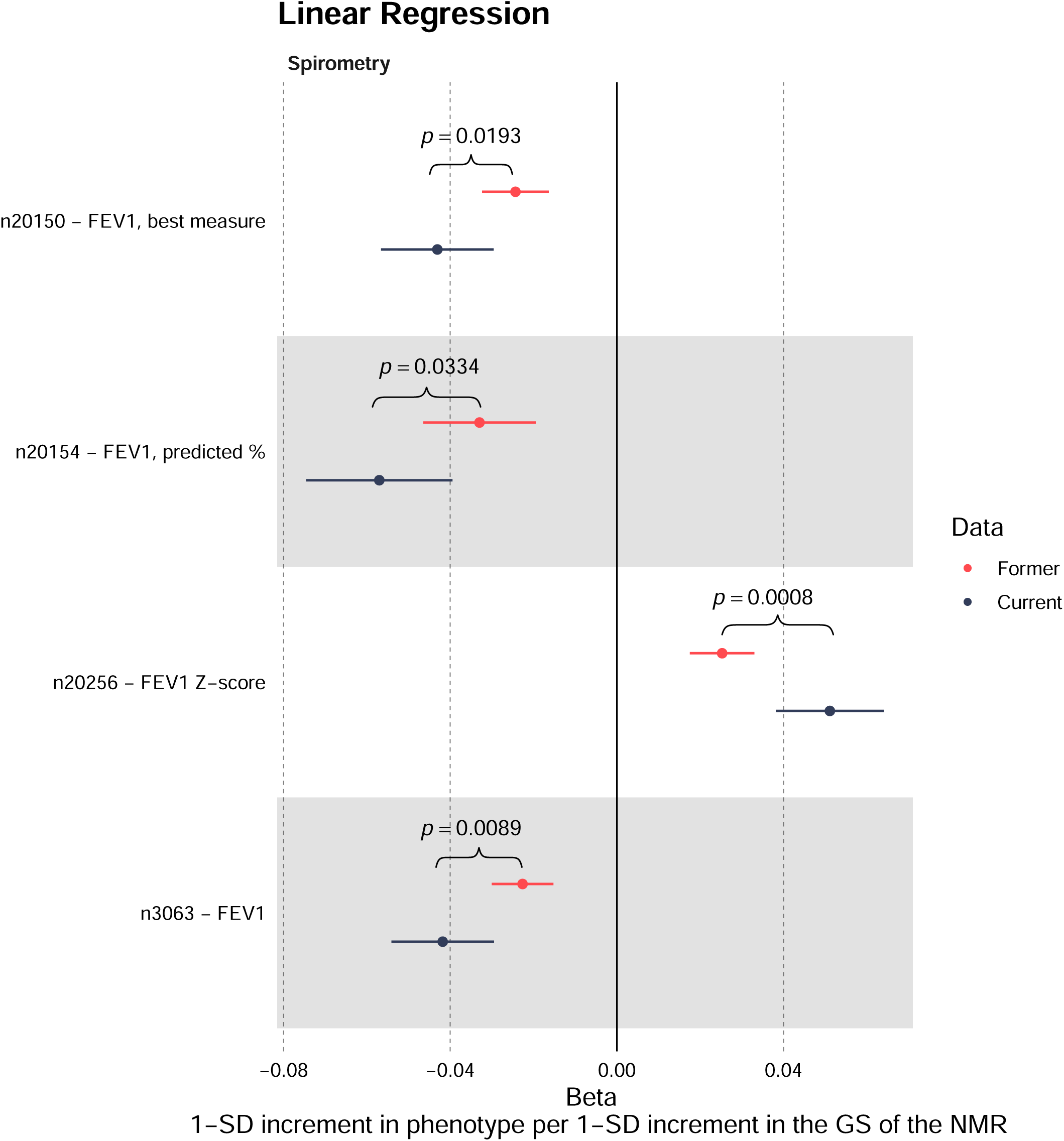
Current versus Former smokers comparison for the four lung capacity variables highlighted in the Ever versus Never smoker analysis. For both Current and Former subsets all four variables had a phenome-wide significant (PWS) association with the GS. Differences between these two subsets were not PWS. Nevertheless, the association is visibly attenuated for the former smokers for all four variables. *n*, normalised after covariates had first been regressed out.

#### GS-Cessation association possibly explained by CPD and ill health

In our sensitivity analyses of cessation, the odds ratio of the GS remained positive after adjusting for CPD (Table S7a–b), although it did decrease from 1.07 (95 % CI: [1.06, 1.09]) to 1.05 [1.04, 1.07]. The odds ratio decreased further to 1.01 [0.97, 1.06] and was no longer statistically significant once we excluded individuals who had stopped due to ‘Illness or ill health’ and/or ‘Doctor’s advice’ (*p* = 0.54) (Table S7c).

When comparing the reasons for stopping smoking (llness or ill health / Doctor’s advice / Health precaution / Financial reasons - participants could choose as many as they wanted), fast metabolizers of nicotine (high GS) seemed to have more incentive to quit than slow metabolizers (low GS), though differences were small (Table S8).

Regardless of reason, fast metabolizers had a slightly higher percentage of people selecting it than slow metabolizers. For the more prevalent reasons: ‘Health precaution’ and ‘Financial reasons’, the differences were statistically significant at the 0.05 level (Table S8).

We took a closer look at ever smokers who had managed to quit at least once for over six months, to see whether any of the reasons for stopping smoking were associated with remaining a former smoker still at the time of the questionnaire (Table S7d–e).

Listing ‘Financial reasons’ was positively associated with remaining a former smoker (OR = 1.21 [1.11, 1.31]), while ‘Doctor’s advice’ had a negative association (OR = 0.61 [0.54, 0.69]) (Table S7d). The GS for faster nicotine metabolism showed a positive association (OR = 1.04 [1.005, 1.08]). After adding CPD to the model, the direction of the association remained the same for all reasons, as well as for the GS (Table S7e).

Now the reasons with p-values below 0.05 were ‘Doctor’s advice’ (OR = 0.51 [0.45, 0.59]), ‘Illness or ill health’ (OR = 0.83 [0.74, 0.94]), and ‘Health precaution’ (OR = 1.08 [1.001, 1.17]). For every additional cigarette the odds of remaining a former smoker was 6 % greater (OR = 1.06 [1.06, 1.07]).

Among former smokers of the Ever subset, the median number of unsuccessful smoking attempts was 2.0, the mean was 2.9 and the standard deviation was 7.1. Once we excluded those listing ‘Illness or ill health’ and/or ‘Doctor’s advice’, the numbers hardly changed: 2.0, 2.8, and 6.9.

All four reasons were statistically significant at the 0.05 level in explaining the number of unsuccessful stop-smoking attempts in the former smokers subset (Table S9). They were all associated with an increased number of attempts, with ‘Health precaution’ demonstrating the strongest association. Individuals listing ‘Health precaution’ had a 39% higher number of attempts compared to those who had not listed it (IRR = 1.39 [1.36, 1.42]) (Table S9a). Additionally, for every standard deviation increase in the GS for the NMR, there was an expected 1 % increase in the number of attempts (IRR = 1.012 [1.002, 1.022]). However, after including CPD in the model, the GS was not statistically significant anymore (*p* = 0.24) (Table S9b). With every additional cigarette, the number of unsuccessful stop-smoking attempts increased by 1.6 % (IRR = 1.016 [1.01, 1.02]).

#### PheWASs in MRBase and FinnGen confirm our findings and highlight lung cancer and lipid outcomes

The NMR top SNP in our previous Meta GWAS of current smokers with European ancestry was rs56113850 on chromosome 19 [15]. Based on our Meta GWAS, the beta for the major allele C (vs. T) was 0.682 (se = 0.02) (C allelle frequency was 0.553). Both the FinnGen and MRBase PheWAS results have been presented for the allele C so that the results can be interpreted for genetically determined faster nicotine metabolism.

The FinnGen PheWAS highlighted two PWS associations: both were for lung cancer diagnoses (Table S10a). The associations were positive, so genetically determined faster nicotine metabolism was associated with greater odds of having lung cancer. The MRBase database pinpointed 199 outcomes with a PWS association (Table S10b).

However, many of these outcomes overlapped or were highly similar (Table S10c). Most of the top associations were related to smoking, lung cancer, lung functioning, liver enzyme levels, cholesterol and other lipids, as well as gene expression (See annotations for highlighted genes: Table S10d). Our own PheWAS of the UKB had not included some of these outcomes such as those related to gene expression, but otherwise the MRBase results closely mirrored ours.

#### Sex and ancestry stratified analyses revealed limited variation

Out of the 71 variables assessed in our sex-stratified analyses, only three showed statistically significant differences between the sexes at a Bonferroni significance level (0.05/71) in at least one of the three data sets (Ever/Never/All) (Fig. S6, Table S12). The genetic score for faster nicotine metabolism was associated with an increased number of cigarettes smoked per day for both sexes but the effect size was greater among males. The GS was negatively associated with glycine. However, this association was absent among male never smokers, while for females, this association was strongest among never smokers. Among current smokers, for smoking compared to 10 years previous (Less nowadays?/About the same?/More nowadays?), there was no association among males but a positive association among females. All variables showing a difference between the sexes that was significant at the 0.05 level have been plotted in Fig. S6, which mainly shows differences in magnitude of effect.

Six of the 33 continuous variables included in our ancestry-stratified analyses were not available for the other ancestry groups. None of the remaining 27 variables showed differences in effect sizes between the ancestry groups and the White British group at a Bonferroni significance level (0.05/27) (Table S13). All variables showing a difference between any of the other ancestry groups and our White British group at the 0.05 level, or showing an effect reaching the 0.05 significance level in one of the other ancestry groups have been plotted in Fig. S7. For both of the liver enzymes, our findings were replicated in some of the other ancestry groups.

## Discussion

In our study we used a hypothesis-free method, GxE MR-PheWAS [22], to identify novel associations across the phenome with the NMR. Importantly, we conducted separate PheWAS analyses for ever and never smokers. This enabled us to identify which associations might be attributable to a causal effect through nicotine metabolism. We explored over 21,000 outcome variables, making our PheWAS the most comprehensive to date on the rate of nicotine metabolism. Notably, our study was the first PheWAS to explicitly focus on the NMR itself, rather than solely on CYP2A6 activity.

We found associations with several smoking related traits and diseases. These were in line with existing literature, with the exception of a positive association between our GS for faster nicotine metabolism and smoking cessation. Additionally, our study unveiled novel associations with measures not previously reported to associate with the NMR, including liver enzymes, lipids, and consumption of coffee and tea.

We did not replicate the hearing loss finding identified by the initial PheWAS of CYP2A6 activity [19]. The other PheWAS published to date on CYP2A6 activity, used the UKB data, but focused only on about 1,000 disease endpoints [20]. They did not replicate the hearing loss finding either. Their findings, limited to tobacco related diseases, were in line with ours, although none of their findings reached phenome-wide significance in our study which included over 21,000 outcome variables. Their top finding, lung cancer, ranked 56th in significance in our subset of ever smokers, for which we saw 29 PWS associations. In our secondary analyses, however, we conducted a PheWAS replication study of the top GWAS SNP for the NMR using the FinnGen data. This FinnGen PheWAS consisted of roughly 2,000 disease endpoints, and here, the only two PWS associations were indeed for lung cancer diagnoses (Table S10a).

Faster nicotine metabolism is known to associate with increased smoking [8, 9]. Our PheWAS results align with this established relationship. The GS for faster nicotine metabolism was associated with a greater number of cigarettes smoked per day, increased smoking compared to ten years earlier, and greater smoking in the past (asked from all except daily current smokers).

We found both the GS and the NMR to have a non-linear association with CPD (Fig 2). This non-linear relationship, indicative of a plateau-effect on CPD, aligns with previous findings for cotinine [41], where a plateau-effect was observed on cotinine when modelling it against CPD. However, to our knowledge, this phenomenon has not been previously reported for the NMR. This relationship provides a possible explanation to why in previous studies *CYP2A6* variation has shown a stronger association with lung cancer risk in light smokers (CPD *≤* 20) than in the total sample [20, 42], and hasn’t shown any association among the heavy smokers (CPD *>* 20) [42]. However, it is possible that, as our population-based data relied on voluntary participation from invitees, some subgroups of individuals with ill health, such as those with high GS and high CPD values, were underrepresented. More research conducted in a controlled setting would be warranted to explore this association more comprehensively.

While clinical trials and longitudinal data have shown that faster nicotine metabolism predicts lower quit rates [11, 12], our PheWAS yielded contrasting results. The GS was associated with greater odds of quitting, and quitting at a younger age (Fig 4). This positive association between genetically determined faster nicotine metabolism and cessation was also seen by Loukola et al. in their follow-up analyses of their NMR GWAS [14]. Cessation was not included in our FinnGen and MRBase replication PheWASs which we performed using the top NMR SNP, rs56113850.

However, the same positive association was seen for the top NMR SNP allele C count with cessation in the GSCAN GWAS [43]. They found that with every additional allele C, the odds for being a former smoker (versus current) increased by 6 % (OR = 1.06 [1.05, 1.07], *p* = 1.61*e −* 48 —calculated from [43] Table S3 to reflect our coding for cessation (1 = former, 0 = current)).

To further investigate the positive relationship between genetically determined faster nicotine metabolism and smoking cessation seen in these population based cross-sectional data, we reran the UKB cessation analyses adding CPD as a covariate to the model. We speculated whether CPD might be acting as a possible mediator for the association. Faster nicotine metabolism is known to be positively associated with CPD. Amount smoked, in turn, impacts one’s health and possibly one’s inclination to quit due to the realization of adverse health effects or the necessity of abstinence, for example, for consequent surgical procedures. After adding CPD into the model, the association weakened but remained positive (Table S7a–b). Once we additionally excluded individuals who listed ill health and/or doctor’s advice as the reason for quitting, the association was no longer statistically significant (Table S7c). Loukola et al. observed similar findings in their follow-up analyses of their NMR GWAS [14].

When we looked at the reasons for quitting more closely, it did appear that fast metabolizers might have had slightly more incentive to quit (Table S8). Additionally, it looked like internal reasons (health precaution and financial reasons) were more likely to result in continued abstinence (remaining a former smoker at the time of the UKB questionnaire) (Table S7d–e). Conversely, having quit because of doctor’s advice seemed to decrease the likelihood of remaining a former smoker at the time of the questionnaire.

As mentioned, those with faster nicotine metabolism are thought to have a harder time succeeding at quitting [10]. Among former smokers, the GS had a positive association with the number of unsuccessful quit attempts but the association was no longer statistically significant once we adjusted for CPD (Table S9). All four reasons increased the number of attempts, as did CPD. Health precaution and ill health however, had considerably stronger associations than doctor’s advice and financial reasons. Our results suggest that several factors are at play when it comes to cessation.

Individuals with faster nicotine metabolism may smoke more, spend more money on tobacco, and experience more health problems. They may thus be more motivated to quit but succeeding can possibly be more challenging due to greater nicotine dependence which has been shown to correlate with faster nicotine metabolism by some studies [42, 44]. Regardless of the difficulties, faster metabolizers of nicotine appear more likely to quit long-term (remain former smokers) than slower metabolizers of nicotine. This could be due to their heightened motivation and more frequent quit attempts. Additionally, due to the nature of the UKB dataset, selection bias may be also contributing to this positive association. The UKB dataset is enriched with individuals who are healthier and come from higher socioeconomic backgrounds [45]. Thus those who smoke more and have not quit may be underrepresented, potentially affecting the results.

In summary, our findings suggest that health precautionary reasons and higher daily cigarette consumption make quitting more challenging. However, they also make it more likely for the individual to remain a former smoker if they do manage to quit. We believe that the positive association between the GS and cessation may be explained, to some extent, by possible confounding or mediation by CPD and ill health, as well as, various selection biases in UKB.

Our Ever versus Never analyses highlighted four lung capacity measures. The associations were only seen in the ever smokers, providing evidence for a causal pathway through nicotine metabolism (Table S6a, Fig 4). When we divided the ever smokers into subsets of former and current smokers, the effect sizes for all four lung capacity measures were attenuated among the former smokers, giving support for the possible causal role of smoking on worse lung functioning (Fig 5, Table S6e). Our results align with existing research. Smoking is known to accelerate the decline in lung function, which may develop into chronic obstructive pulmonary disease [46, 47]. It is worth noting that, although smoking cessation has been shown to slow down this rate of decline, it does not appear to fully revert the rate of decline to that of never smokers [48].

Beyond variables related to smoking, our study identified several other variables associated with our GS for the NMR. It is noteworthy that many of these associations did not seem to rely on pathways through nicotine metabolism. Interesting associations, where we saw no apparent differences between ever and never smokers, included liver enzymes, lipid measures, and coffee and tea consumption.

In our PheWAS, only two phenotypes, the liver enzymes alkaline phosphatase (ALP) and alanine aminotransferase (ALT), showed associations with the GS that were PWS in both ever and never smokers (Fig 4). These associations ranked among the top findings in all three groups (All, Ever, Never), as well as in the PheWAS results from MRBase (Table S6b–d, Table S10b).

Both ALP and ALT are blood serum liver enzymes that are key biomarkers for assessing the extent and cause of liver damage [49–51]. One cause of liver damage is alcohol use, and alcohol use and smoking are well known to be associated behaviours. However, the strengths of the associations did not differ between the ever and never smokers, implying that the associations are not mediated by nicotine metabolism or smoking, and thus most likely not by alcohol either.

Building upon previous PheWAS analyses of these liver enzymes, conducted by Liu et al. [49], our study suggests that a higher GS for faster nicotine metabolism is correlated with less favourable liver enzyme levels with respect to associated diseases. We observed a negative association between the GS and ALP levels. The ALP PheWAS by Liu et al., revealed that lower values of the genetically determined ALP were associated with increased odds of hypercholesterolemia, pulmonary heart disease, as well as phlebitis and thrombophlebitis of lower extremities.

On the other hand, we found a positive association between the GS and ALT levels. For genetically determined ALT levels, Liu et al. found 16 associations in their ALT PheWAS. The strongest associations, based on p-values, highlighted a clear trend: higher values of genetically determined ALT levels were consistently associated with a higher risk of hepatic diseases such as hepatitis, primary liver cancer and non-alcoholic cirrhosis. Additionally, genetically determined ALT was positively associated with Type 2 Diabetes, possibly due to the role ALT plays in insulin resistance [49]. In our PheWAS of the entire sample, we identified a PWS positive association between the GS and glycated haemoglobin, suggesting that higher GS values correlate with poorer blood sugar control. The effect size remained consistent across the Ever and Never subsets but did not reach PWS in ether subset. Of the 16 associations reported by Liu et al. for ALT, only three showed an opposite direction of effect: dementias, fracture of hand and wrist, and corneal degenerations. However, all three had p-values that had only just surpassed the 5 % FDR threshold, and for dementias, the direction of the effect varied depending on the Mendelian Randomization analysis method they used.

A substantial portion of our MRBase PheWAS results concerned outcome variables related to cholesterol, fatty acids, and other lipids (Table S10c). Likewise, other outcomes indicative of cardiovascular health were also highlighted. For instance, genetically determined faster nicotine metabolism (the NMR top SNP, rs56113850, allele C count), was associated with elevated levels of cholesterol, triglycerides, LDL levels, and a higher apolipoprotein B/apolipoprotein A ratio (indicative of poorer cardiovascular health), as well as an increased waist circumference, higher BMI and greater odds of using cholesterol medication. Of note, cholesterol medication was among the top five associations in the PheWAS of the FinnGen data, based on p-values, although it did not reach phenome-wide significance.

In our UKB PheWAS, the GS was associated with decreased apolipoprotein A, increased linoleic acid, a higher omega-6 to omega-3 ratio and a decreased omega-3 to total fatty acids ratio (Fig 4). Again, the direction of association was such that higher GS values predicted worse lipid values with respect to cardiovascular health (see e.g. [52]). These results were in line with our findings from the MRBase PheWAS. However, they were PWS only in our All group. Nevertheless, there was no apparent difference between the effect size estimates among the ever and never smokers, alluding to effects independent of nicotine metabolism, and possibly to a shared genetic component between nicotine metabolism and lipid levels. Interestingly, only among ever smokers, we observed PWS positive associations between the GS and the use of cholesterol medication, leg artery bypass surgery, and endarterectomy of femoral artery (Fig 4). This suggests that ever smokers with higher GS values may face a compounded risk for cardiovascular diseases due to their smoking history and genetics, and may thus be more likely to face adverse health effects and require medical interventions.

Moving on to another noteworthy association highlighted in our PheWAS, the GS for faster nicotine metabolism was associated with increased coffee and tea consumption (Fig 4). We observed PWS associations with coffee consumption in the All group and with tea consumption in the All and Never groups. The concordant results between Ever and Never subsets suggest an association pathway distinct from nicotine metabolism.

When decaffeinated coffee was included in the coffee quantity variable, there was a slight attenuation of associations, suggesting that the observed effect is possibly driven by caffeine metabolism. Notably, in a GWAS of caffeine metabolites, multiple SNPs at 19q13.2, including the *CYP2* cluster, *NUMBL*, *ADCK4*, *MIA* and *EGLN2*, showed genome wide significant associations [53]. The CYP2A6 enzyme is known to play a minor role in caffeine metabolism [53], and it is possible that our results reflect this connection.

Interestingly, our PheWAS also highlighted a positive correlation between the GS and preferences for dark chocolate and coffee without sugar. These associations were PWS only in the All subset. However, both the Ever and Never results were consistent with those of the All subset. Once again, this would imply that the association pathway is unlikely to involve nicotine metabolism. Like coffee and tea, dark chocolate contains caffeine and tastes bitter. Therefore, these findings suggest that the association with coffee and tea consumption could be mediated through preference for bitter taste in addition to caffeine metabolism. Cornelis and van Dam (2021) found in their study that genetically inferred bitter taste perception indeed did play a role in coffee and tea drinking behaviour but to a lesser extent than genetically inferred caffeine sensitivity [54]. They also found support for conditioned taste preferences where individuals learn to associate the bitter taste with either beneficial or adverse physiological effects of caffeine.

While we did not observe direct evidence of a pathway involving smoking, it is well known that smoking and coffee consumption correlate [55]. The prevailing belief is that tobacco smoke increases an individual’s caffeine metabolism and that is why smokers require more caffeine to achieve the desired effect [56]. Once an individual stops smoking, their caffeine metabolism eventually returns to normal. A previous Mendelian randomization study found support for this relationship [57]. They compared current, former and never smokers using a single SNP as a biomarker for CPD [57]. They found no association with coffee consumption among never or former smokers in their meta-analysis of three datasets but did find an association among current smokers.

Our results align with the work of Cornelis and van Dam, suggesting some shared genetics between nicotine metabolism and caffeine metabolism, along with potential connections to bitter taste perception. Both smoking and caffeine impact the metabolism of some medications and affect optimal dosages [56]. Additionally, it has been proposed that smokers attempting to quit, may inadvertently confuse symptoms of caffeine toxicity with nicotine withdrawal symptoms if they fail to adjust their coffee consumption [55]. Thus, understanding the interactions between smoking and caffeine consumption is of great interest.

The main limitation of our study was the potential bias due to UKB’s enrichment with healthier individuals. Additionally, we had limited statistical power to study ancestry groups other than the White British group, as the sample sizes are notably smaller. This limits the accuracy of the effect size estimates for the other groups, and thus our power to detect differences with the White British group or to observe replication of our results. Based on our ancestry-stratified follow-up analyses, the other ancestry groups did not seem to notably differ from our White British sample, admittedly possibly due to a lack of statistical power. We were, however, able to replicate our liver enzyme findings in some of the other ancestry groups. Another limitation is that there may be some overlap between the individuals included in our NMR GWAS and those analyzed in the top NMR SNP (rs56113850) PheWASs from MRBase and FinnGen. Considering that the GWAS and Sequencing Consortium of Alcohol and Nicotine use (GSCAN) (n = 243,952) is the most likely cohort in MRBase to have overlap with our data, and that the total sample size for FinnGen was 377,277, our NMR GWAS was conducted with a relatively small sample size of 5,185 individuals (of whom 2,572 were from Finland). Thus, we expect that the overlapping individuals have minimal impact on the estimated effect sizes for most of the outcome variables in the FinnGen and MRBase PheWASs. A further limitation, as discussed by Buchwald et al. [15], was that individual-level genotype and NMR data required for the fine-mapping analyses of the NMR loci were only available from two Finnish datasets. Thus, the weights used for the chromosome 19 SNPs in the genetic score rely solely on these datasets. However, one of our main strengths was the ability to perform FINEMAP analysis: the variance explained by the chromosome 19 FINEMAP top configuration was 32.1 % as opposed to 23 % explained by the top SNP alone. Additionally, apart from having a very strong instrumental variable, another main strength of our study was the implementation of the GxE MR approach, which involved comparing ever versus never smokers and allowed for inferences about the association pathways.

## Conclusions

This study presents the first comprehensive PheWAS of the NMR, uncovering novel associations not previously reported in the context of nicotine metabolism. Our results suggest that genetically determined faster nicotine metabolism is associated not only with smoking related traits but also with various adverse health outcomes. Our GS for faster nicotine metabolism was associated with worse liver enzyme and lipid values with respect to associated diseases, as well as increased coffee and tea consumption.

Importantly, we saw no evidence of a causal pathway through nicotine metabolism for these associations. Leaning on the assumption that slow metabolizers of nicotine have an easier time quitting, our findings are promising. They support a possibility that a future smoking cessation therapy targetting genes associated with the NMR, to convert fast metabolizers into slower metabolizers, could work without adverse side effects and potentially even provide other health-related benefits. Future research, focusing on these newly highlighted variables using additional data and study designs, involving different sources of potential bias, is warranted to confirm and extend these findings.

## Availability of data and materials

Access to the UK Biobank data can be applied at http://ukbiobank.ac.uk/register-apply/. To apply for access to the YFS data see: http://youngfinnsstudy.utu.fi/, and for FINRISK see: https://thl.fi/en/web/thl-biobank. All analyses were performed in R and bash. Scripts will be deposited on GitHub upon publication of the manuscript.

## Supporting information

**Fig. S1 Pipeline plot summarising the methods and UKB data used.** Main analyses are in pink and follow-up analyses in blue.

**Fig. S2 Boxplots of the standardised GS for the NMR in UKB by subgroups.** Experimenters includes those individuals who answered “Occasionally” or “Tried once or twice” to the questions on current and past smoking behaviour.

**Fig. S3** Scatterplot of the GS for faster nicotine metabolism against the imputed genotype dosage (allele C) at the chromosome 19 top SNP (rs56113850) for the NMR in UKB.

**Fig. S4 Scatterplots and loess curves presenting the full data of the association between A** the standardized genetic score for the NMR (zGS) and cigarettes smoked per day (CPD) in UKB, **B** the standardized NMR (zNMR) and CPD in the Finnish data, **C** the NMR and CPD in the Finnish data, and **D** the zNMR and Cotinine + 3-Hydroxycotinine (Cot + 3HC), a biomarker for nicotine intake, in the Finnish data. All plots are for current smokers. The standardized variables (zGS and zNMR) were calculated by subtracting the mean and dividing by the standard deviation.

**Fig. S5 Venn diagram of the 61 variables highlighted in our initial PheWAS.** The figure shows the 61 variables that were statistically significant at the 5% FDR level in at least one of the data sets (All / Ever / Never). The variable Smoking status has been listed twice in the figure as it contained a different amount of categories for the All and Ever groups.

**Fig. S6 Forest plot of the 18 variables highlighted in our sex stratified analyses of the 71 variables that were included in our final PheWAS.** Results for males have been indicated with squares and results for females with diamonds. Solid circles/squares indicate a statistically significant effect size at *p <* 0.05. The figure shows the variables that had a statistically significant (*p <* 0.05) difference between the effect sizes of females and males in at least one of the data groups (Never/Ever/All). *, there was a bonferroni significant (*p <* 0.05*/*71) difference between the effect sizes of the males and females, *n*, normalised after covariates had first been regressed out; *d*, derived from the original UKB phenotype; *c*, coding corrected to be more intuitive.

**Fig. S7 Forest plot of the 11 variables highlighted in our ancestry-stratified analysis of the 33 continuous variables that were phenome-wide significant in our final PheWAS.** The figure shows the 11 variables that were statistically significant (p ¡ 0.05) in at least one of the ancestry groups (solid circles/squares), other than White British, or had a statistically significant difference in their effect sizes as compared to the White British group (p ¡ 0.05) (square shape). *n*, normalised after covariates had first been regressed out; *d*, derived from the original UKB phenotype.

**Table S1 GS distribution by subset of smoking status.** The p-values are from Mann-Whitney U tests comparing the GS distributions to the Never group. *sd*, standard deviation.

**Table S2 FINEMAP top configuration of causal SNPs for the NMR on the chromosome 19 locus in the Finnish data.** FINEMAP results of the 5Mb region centred at the top SNP when only including SNPs passing quality control in UKB (n(SNP) = 10,133). The most probable configuration consisted of 9 SNPs (depicted in the table) and their heritability estimate was 32.1% (95% CI: 28.5–35.6%). FINEMAP gave a regional heritability estimate of 33.8% (95% CI: 30.0–37.8%) and suggested that there are 7–11 causal SNPs within the region. *SNP*, single-nucleotide polymorphism; *BP*, base pair position in GRCh37 coordinates; *EA/NEA*, the effect allele/ the non-effect allele; *MINOR*, the less common allele; *MAF*, minor allele frequency in the Finnish dataset (n = 2,119) used for the FINEMAP analysis; *SNP PROB*, posterior probability of being a causal SNP; *BETAJ*, effect estimate from the joint model including all these 9 SNPs (reported for the effect allele); *SEJ*, standard error for the BETAJ; *P-VALUEJ*, p-value for the BETAJ.

**Table S3 Variable descriptions for PheWAS outcomes. A** Variable information on all the second stage outcomes that we derived, recoded or analysed using a different model from our initial PheWAS (See Table S3b). **B** Variable information on all 61 variables highlighted in our initial PheWAS analyses.

**Table S4 Linear regression beta coefficient for the zGS when explaining CPD by zGS in UK Biobank.** Analysis was done for the current smokers subset of UK Biobank. CPD has been adjusted for sex, age and the first 10 genetic principal components, and then inverse normalized (rank-based-inverse-normal-transformation) before regressing it on zGS. The standard deviation of the adjusted CPD was 8.3. When looking at all individuals, each standard deviation increase in the GS is associated with a 0.07516 standard deviation increase in CPD, i.e. a 0.6 increase in cigarettes smoked per day. The regression was ran for three subsets of the data including either all individuals (ALL), only those with lower zGS values (LOW GS; zGS *<* 0) or only those with higher zGS values (HIGH GS; zGS *≥* 0).

**Table S5 Results from the initial PheWAS. A-C** PheWAS results for the variables that were significant at the 5 % FDR level for the All, Ever and Never data, respectively. **D** Ever versus Never analysis results for the variables that had a statistically significant difference at the 5 % FDR level between their effect sizes for the Ever and Never subsets.

**Table S6 Results from the final stage of the PheWAS. A** Ever versus Never analysis results. **B-D** PheWAS results for the All, Ever and Never data, respectively. **E** Current versus Former analysis results for the four lung capacity measures that had been highlighted in the Ever versus Never analysis (See Table S6a).

**Table S7 Odds ratios from the logistic regression model for cessation. A** Among Ever smokers (n = 110348), model including the standardized GS, sex (1=Male, 0=Female), age and the first 10 genetic principal components as the predictor variables. **B** Among Ever smokers (n = 110348), model including CPD as an additional predictor variable. **C** Among subset of Ever smokers who did not stop smoking due to Illness or Doctor’s advice (n = 70278), model including all the same variables as in B. **D** Among subset of Ever smokers who had at least once managed to quit for over 6 months (n = 83704), model including the 4 reasons for stopping smoking. **E** Among subset of Ever smokers who had at least once managed to quit for over 6 months (n = 79228), model including the 4 reasons for stopping smoking and CPD.

**Table S8 Reason for stopping smoking by nicotine metabolism group (grouped based on the tertiles of the GS for the NMR).** Ever smokers who had stopped smoking for over 6 months during the time they smoked were asked: “Why did you stop smoking? (You can select more than one answer)” (UK Field 6157). Total number of ever smokers included in this analysis was n = 110 348, of which 26 % were current smokers and 74 % former smokers. *, statistically significant difference (p *<* 0.05, 2-sample test for equality of proportions) between the Slow and Fast groups.

**Table S9 Incident Rate Ratios (IRRs) from the negative binomial regression model for Number of unsuccessful stop-smoking attempts. A** Among former smokers, model including standardized GS, the four reasons (1 = Yes, 0 = No), sex (1 = Male, 0 = Female), age and the first 10 genetic principal components as the predictor variables. **B** Same as A but including CPD as a covariate in the model.

**Table S10 The FDR significant results from the PheWAS of the top NMR SNP, rs56113850 (allele C), using FinnGen and MRBase. A** Using the FinnGen r9 there were two outcomes reaching statistical significance at the 0.05 FDR level. Beta has been counted for the C allele. Allele C frequency (vs T) was 56–57 % across all phenotypes. *mlogp*,-log10(p); *Bhcritical*, Benjamini-Hochberg critical value. **B** Using the MRBase (Database version: 0.3.0 from 25 October 2020) there were 199 outcomes reaching statistical significance at the 0.05 FDR level. *minuslogp*,-log10(p); *Bhcritical*, Benjamini-Hochberg critical value; *bf*, significant at the 0.05 bonferroni level (T=TRUE, F=FALSE). **C** Same as B but ordered by trait. **D** Annotations for the 14 gene expression outcomes that were among the 199 MRBase results. Annotations were obtained from the Ensembl database using BiomaRt in R. *band*, Karyotype band; *gene biotype*, Gene type; *hgnc symbol*, HGNC (The HUGO Gene Nomenclature Committee) gene symbol. Note: MRBase id column can be searched as the GWAS ID here: https://gwas.mrcieu.ac.uk/datasets/ for more information on the study.

**Table S11** Information on the NMR top SNP, rs56113850, and smoking status by ancestry group.

**Table S12 Sex-stratified analyses.** Results for all 71 variables assessed.

**Table S13 Ancestry-stratified analyses.** Results for all 33 continuous variables assessed.

## Acknowledgments

This research has been conducted using the UK Biobank Resource under Application Number 22627. We thank Dr. Samuel Jones for organizing the UK Biobank data files. We want to express our gratitude to all participants in UK Biobank, YFS, FINRISK and FinnGen as well as to all people, institutes and funding bodies that have been involved in the infrastructure surrounding these cohorts, and made these valuable resources possible.

## Author contributions

Jadwiga Buchwald: Conceived and designed the study; acquired funding; determined the statistical analysis methodology; curated data; conducted the analysis; interpreted and visualized the data; wrote the manuscript. Matti Pirinen: Conceived and designed the study; acquired funding; determined the statistical analysis methodology; curated data; interpreted the data; supervised the project; critically reviewed and revised the manuscript. Jaakko Kaprio: Conceived and designed the study; acquired funding; critically reviewed and revised the manuscript. Terho Lehtimäki: Acquired data. Olli Raitakari: Acquired data. Veikko Salomaa: Acquired data. All authors read and approved the final version of the manuscript and agree to be accountable for all aspects of the work.

## Ethics approval and consent to participate

UKB was approved by the North West Multi-center Research Ethics Committee. The Coordinating Ethics Committee of the Helsinki and Uusimaa Hospital District approved the FINRISK surveys (decision number 229/E0/06 in 2007 and 162/13/03/00/11 in 2012). The local University Hospital ethics committees in Turku and Tampere approved the YFS study. All participants provided written informed consent.

