## Supplementary Information for "A Phenome-wide association study of genetically determined nicotine metabolism reveals novel links with health-related outcomes"

### Contents

**1 Supplementary Figures** **3**

**2 Supplementary Tables** **12**

**3 Extended Content** **15**

3.1 FINEMAP analyses . . . . . . . . . . . . . . . . . . . . . . . . . . . . . . . . . . . . . 15

3.2 The GxE MR-pheWAS approach . . . . . . . . . . . . . . . . . . . . . . . . . . . . . . 15

### 1 Supplementary Figures

4 **Fig S4.** Scatterplots and loess curves presenting the full data of the association between **(A)** the standardized genetic score for the NMR (zGS) and cigarettes smoked per day (CPD) in UKB, **(B)** the standardized NMR (zNMR) and CPD in the Finnish data, **(C)** the NMR and CPD in the Finnish data, and **(D)** the zNMR and Cotinine + 3-Hydroxycotinine (Cot + 3HC), a biomarker for nicotine intake, in the Finnish data. All plots are for current smokers. The standardized variables (zGS and zNMR) were calculated by subtracting the mean and dividing by the standard deviation. . . . . . . . . . . . . . . . . . . . . . . . 6

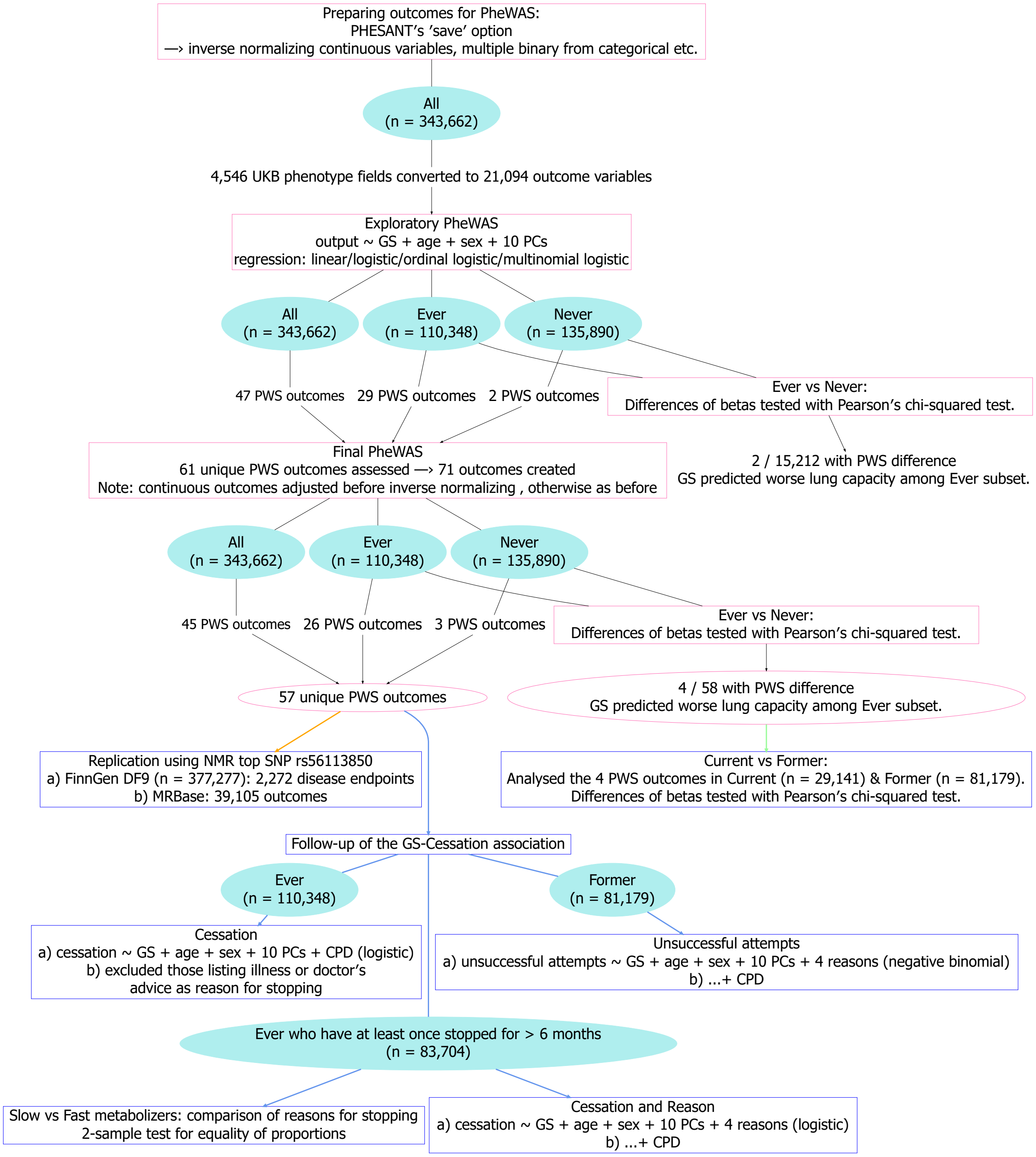

**Fig S1.** Pipeline plot summarising the majority of methods and UKB data used. Main analyses are in pink and follow-up analyses in blue. *GS*, Genetic Score; *PWS*, Phenome-wide significant; *NMR*, Nicotine Metabolite Ratio; *CPD*, Cigarettes smoked per day; *10 PCs*, first ten principal components of genetic structure.

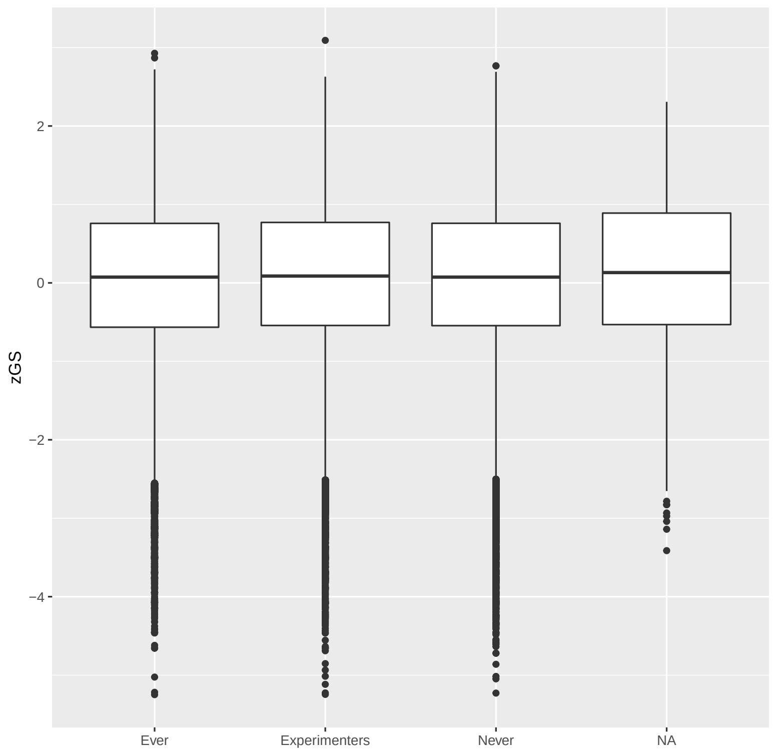

**Fig S2.** Boxplots of the standardised GS for the NMR in UKB by subgroups. Experimenters includes those individuals who answered "Occasionally" or "Tried once or twice" to the questions on current and past smoking behaviour.

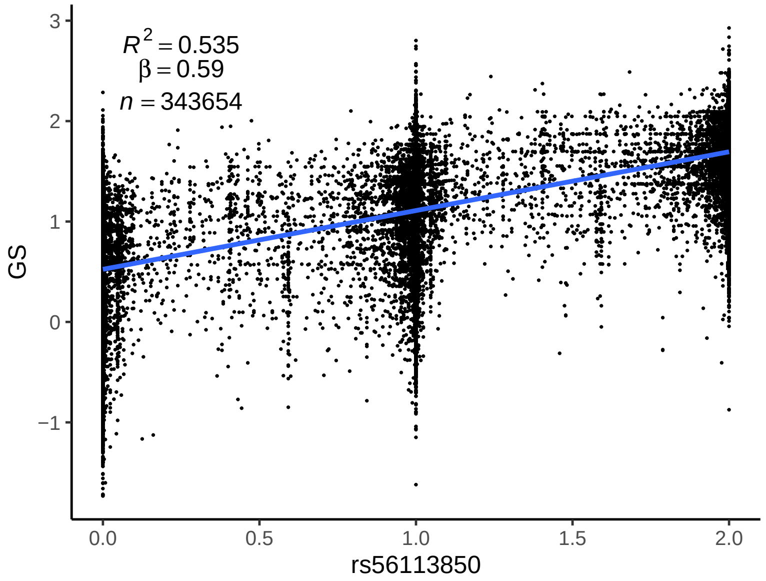

**Fig S3.** Scatterplot of the GS for faster nicotine metabolism against the imputed genotype dosage (allele C) at the chromosome 19 top SNP (rs56113850) for the NMR in UKB.

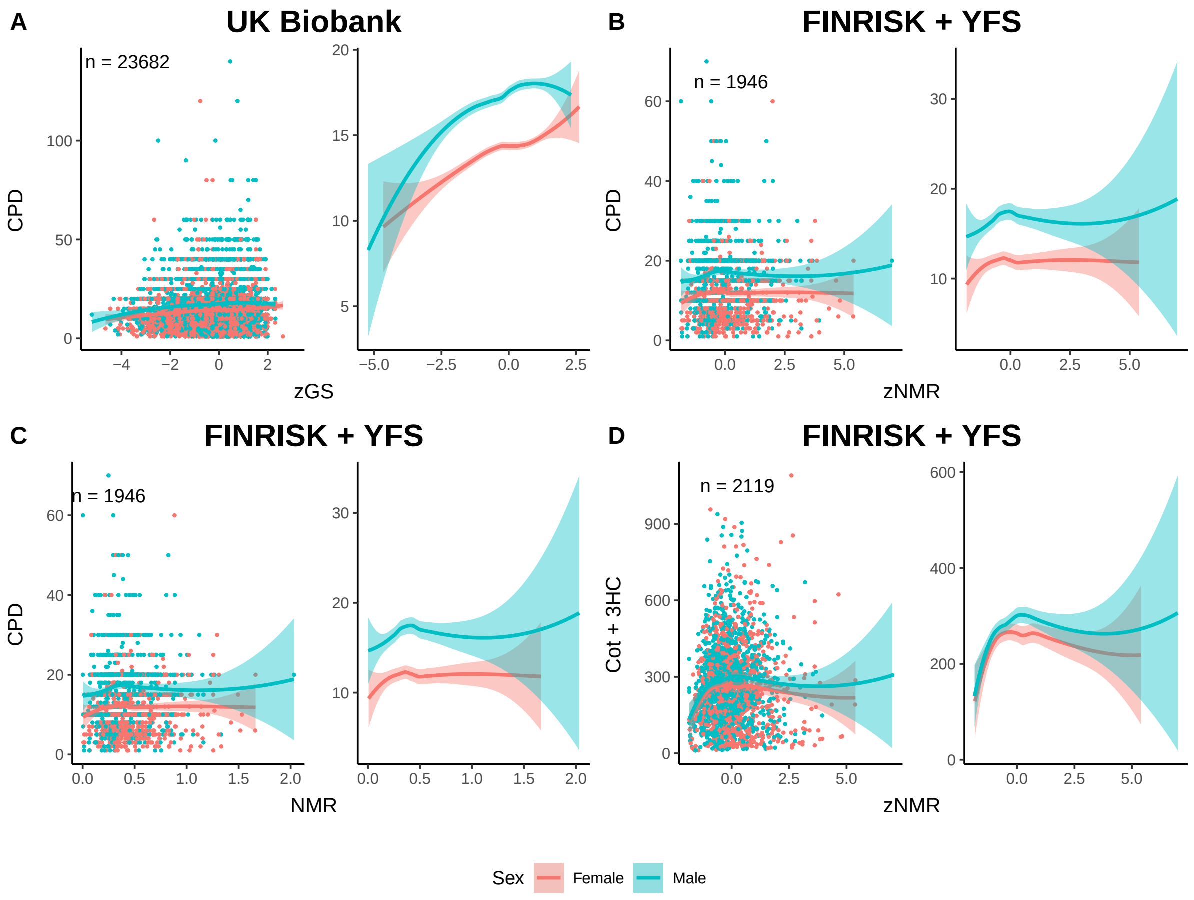

**Fig S4.** Scatterplots and loess curves presenting the full data of the association between **(A)** the standardized genetic score for the NMR (zGS) and cigarettes smoked per day (CPD) in UKB, **(B)** the standardized NMR (zNMR) and CPD in the Finnish data, **(C)** the NMR and CPD in the Finnish data, and **(D)** the zNMR and Cotinine + 3-Hydroxycotinine (Cot + 3HC), a biomarker for nicotine intake, in the Finnish data. All plots are for current smokers. The standardized variables (zGS and zNMR) were calculated by subtracting the mean and dividing by the standard deviation.

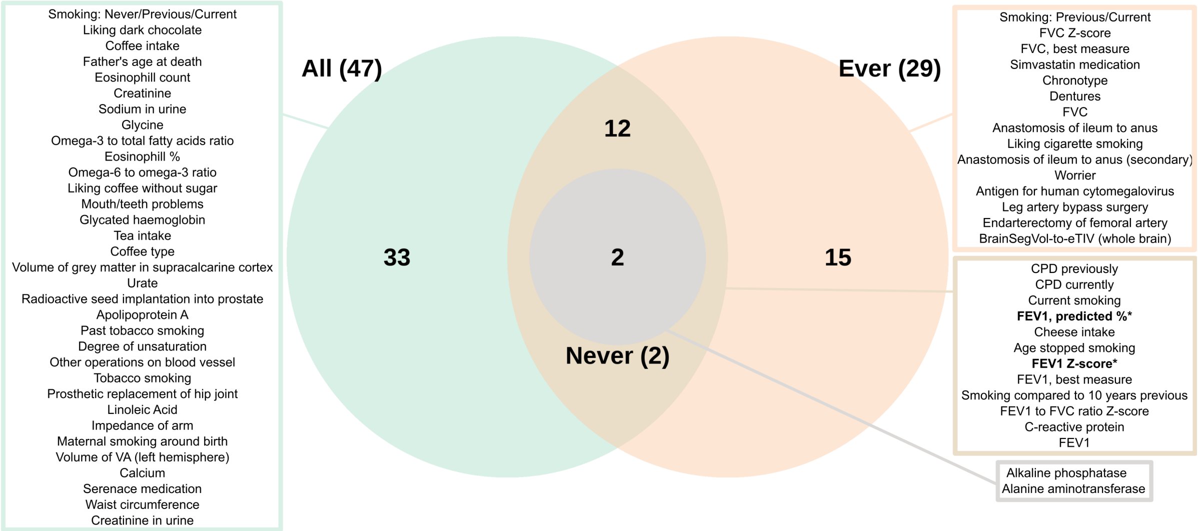

**Fig S5.** Venn diagram of the 61 variables highlighted in our initial PheWAS. The figure shows the 61 variables that were statistically significant at the 5 % FDR level in at least one of the data sets (All / Ever / Never). The variable Smoking status has been listed twice in the figure as it contained a different amount of categories for the All and Ever groups.

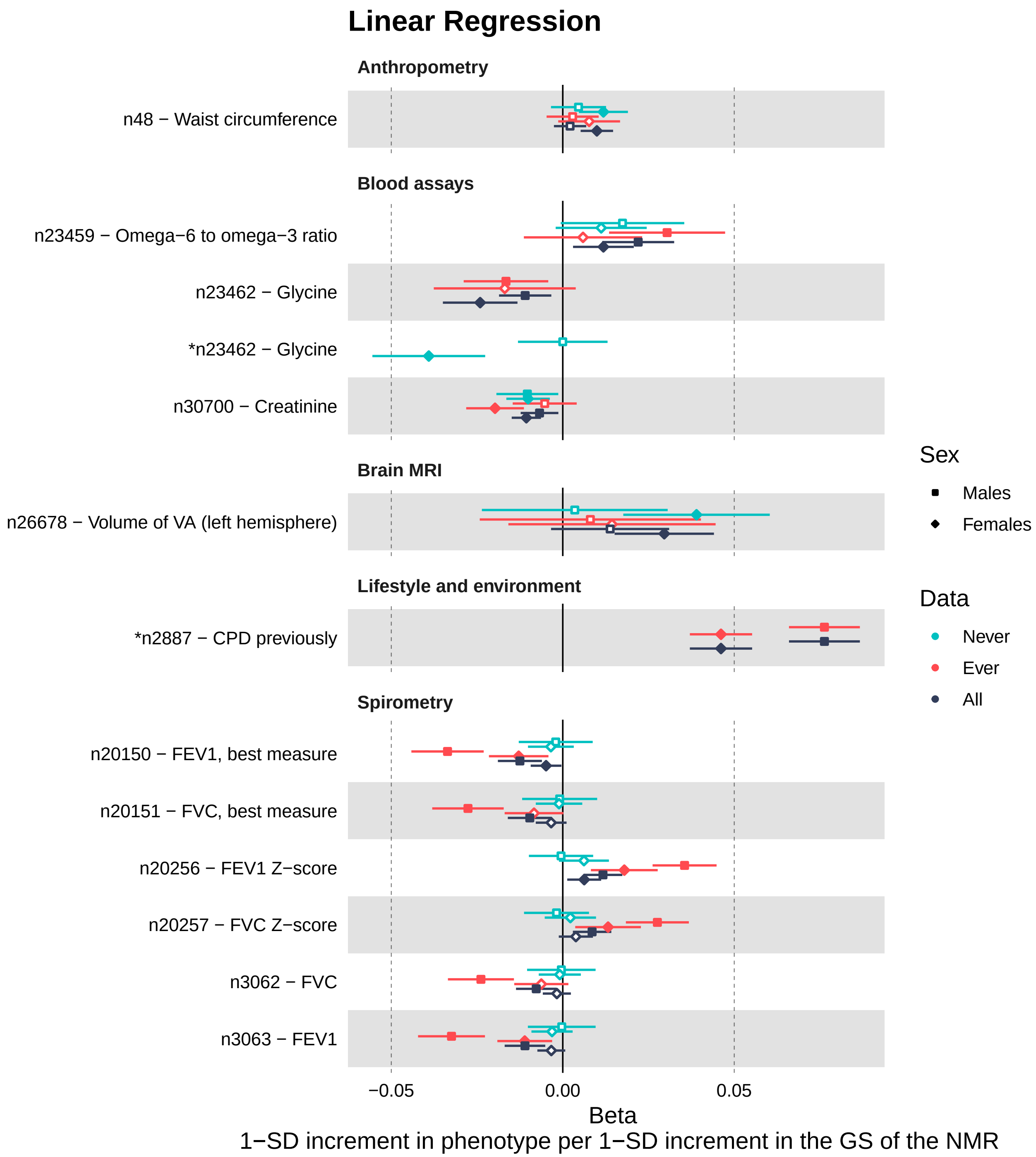

**Fig S6.** Forest plot of the 18 variables highlighted in our sex-stratified analyses of the 71 variables that were included in our final PheWAS. Results for males have been indicated with squares and results for females with diamonds. Solid circles/squares indicate a statistically significant effect size at $p < 0.05$. The figure shows the variables that had a statistically significant ($p < 0.05$) difference between the effect sizes of females and males in at least one of the data groups (Never/Ever/All). *, there was a Bonferroni significant ($p < 0.05/71$) difference between the effect sizes of the males and females; $n$, normalised after covariates had first been regressed out; $d$, derived from the original UKB phenotype; $c$, coding corrected to be more intuitive.

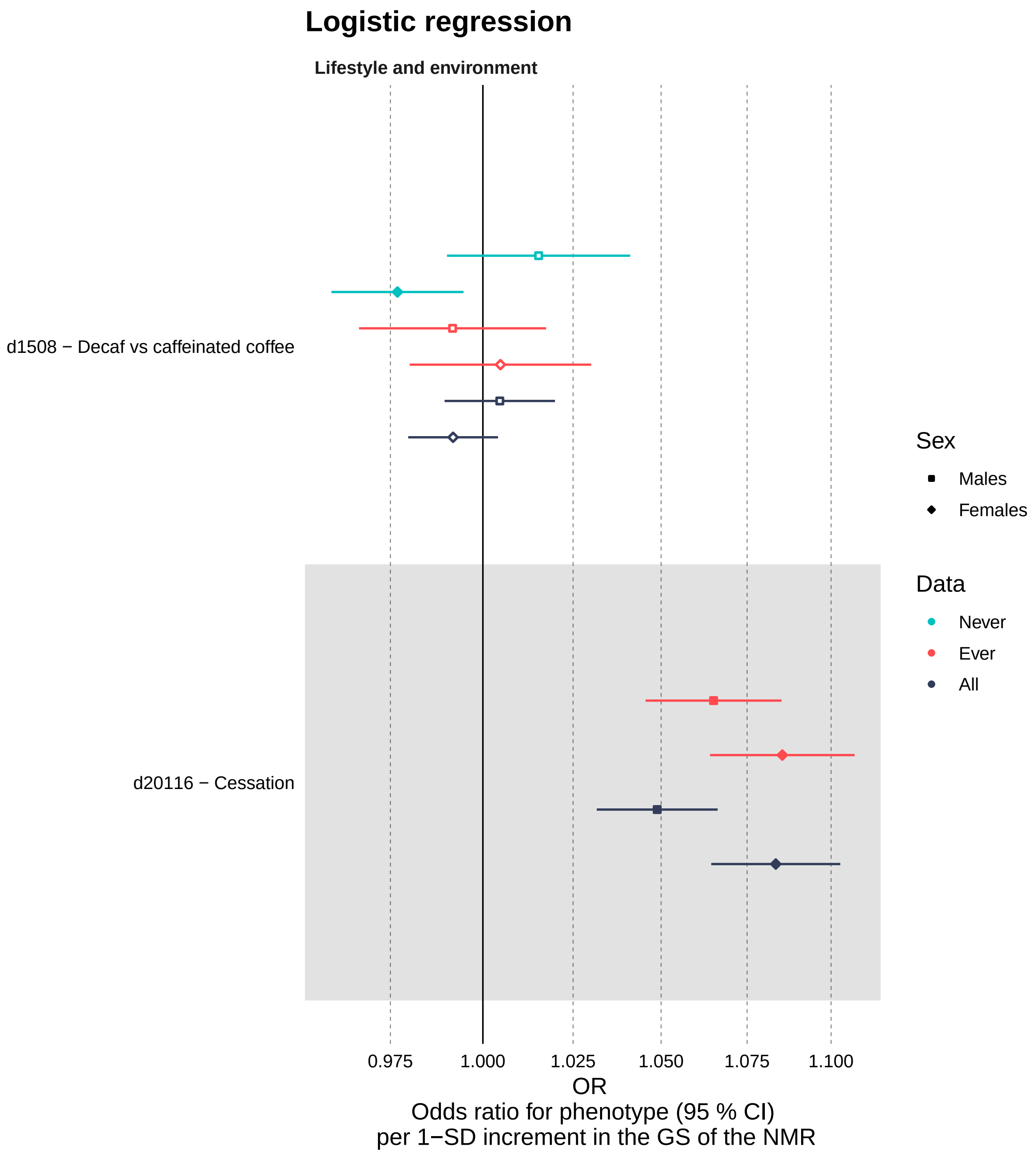

**Fig S6** continued

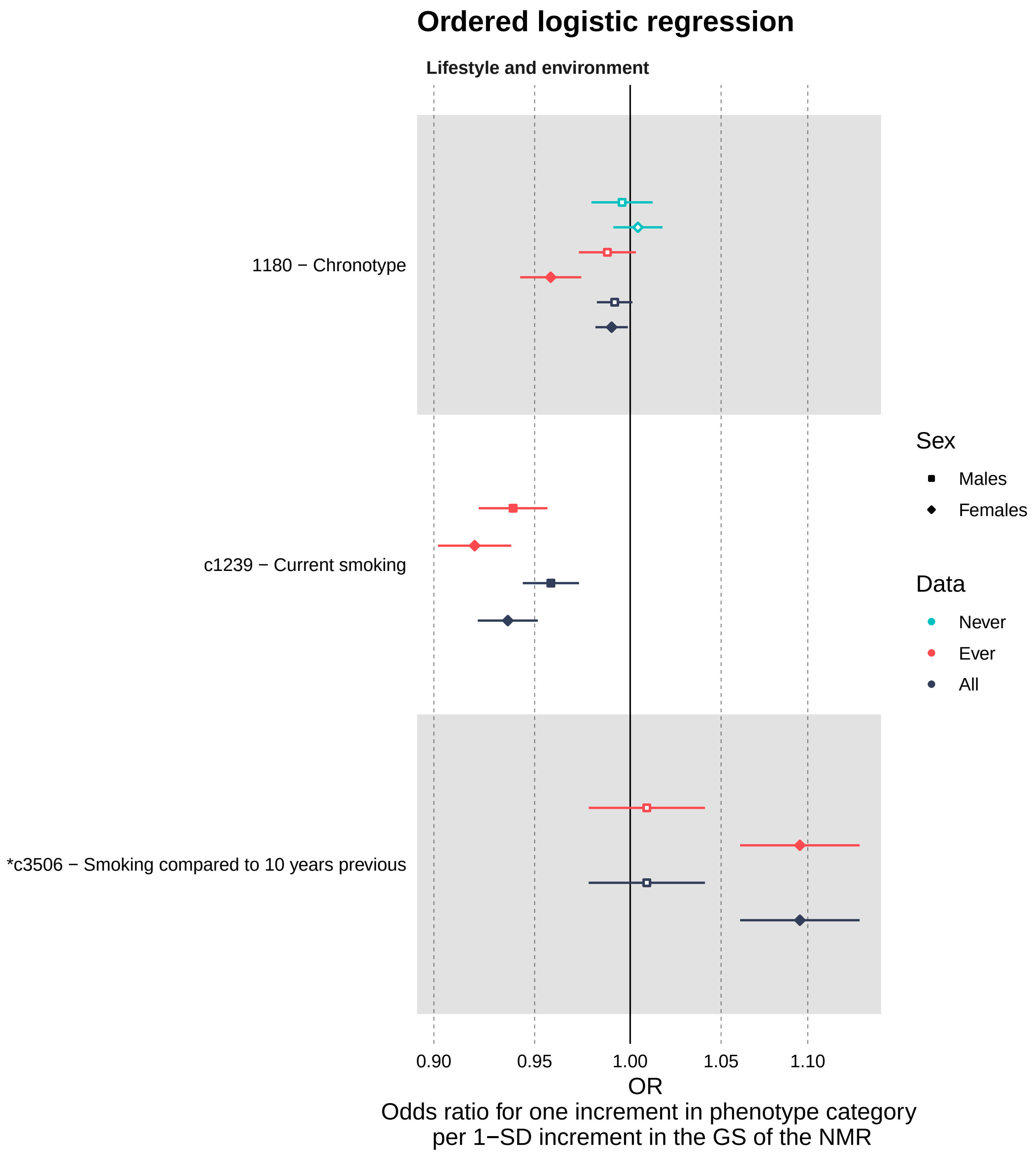

**Fig S6** continued

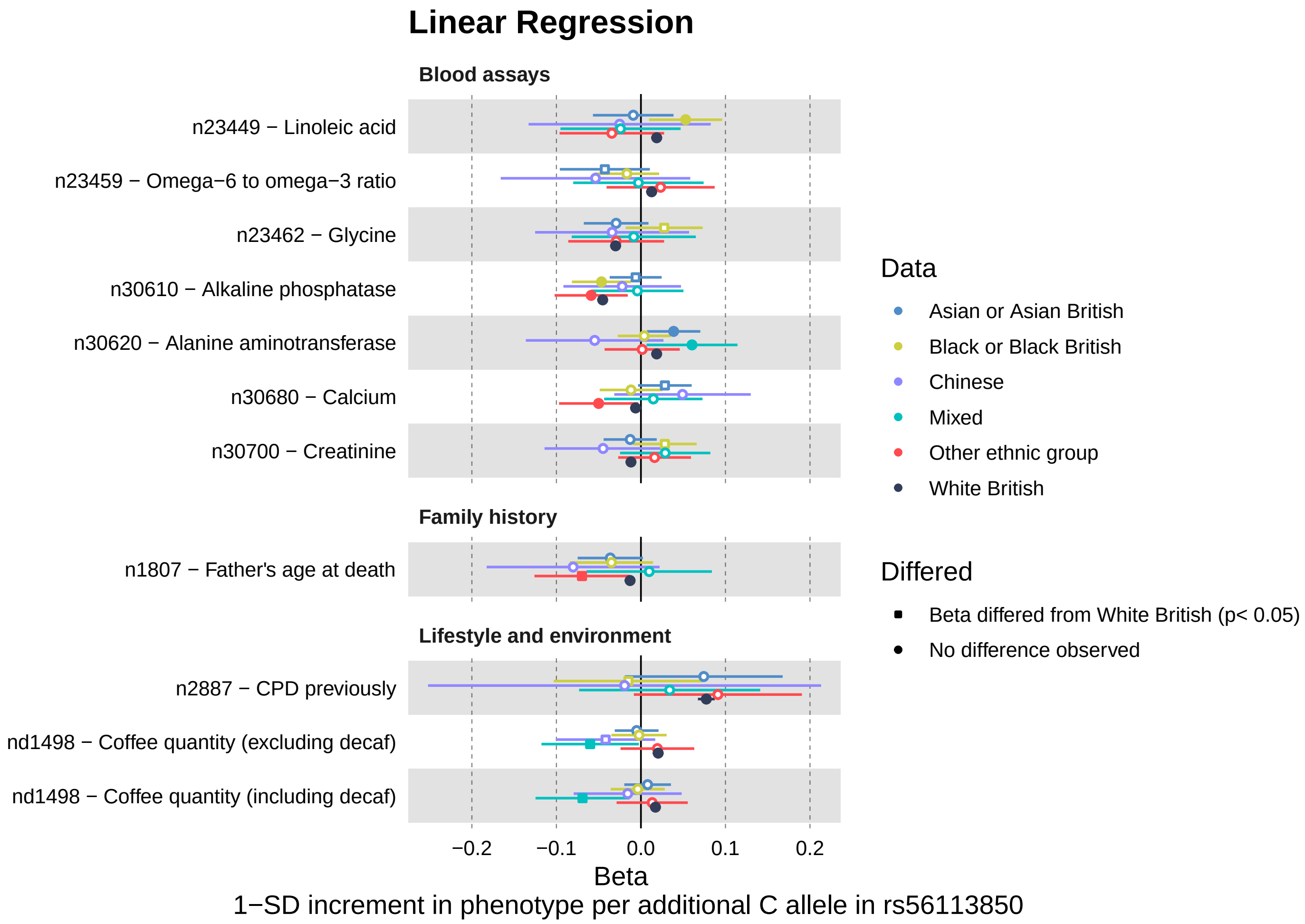

**Fig S7.** Forest plot of the 11 variables highlighted in our ancestry-stratified analysis of the 33 continuous variables that were phenome-wide significant in our final PheWAS. The figure shows the 11 variables that were statistically significant ($p < 0.05$) in at least one of the ancestry groups (solid circles/squares), other than White British, or had a statistically significant difference in their effect sizes as compared to the White British group ($p < 0.05$) (square shape). $n$, normalised after covariates had first been regressed out; $d$, derived from the original UKB phenotype.

### 2 Supplementary Tables

#### List of Excel Tables Provided Separately

Supplementary Tables 3, 5, 6, 7, 9, 10, 12 & 13 are provided as excel tables seperately.

**Table S3 Variable descriptions for PheWAS outcomes. A** Variable information on all the second stage outcomes that we derived, recoded or analysed using a different model from our initial PheWAS (See Table S3b). **B** Variable information on all 61 variables highlighted in our initial PheWAS analyses.

**Table S12 Sex-stratified analyses.** Results for all 71 variables assessed.

**Table S13 Ancestry-stratified analyses.** Results for all 33 continuous variables assessed.

**Table S1.** GS distribution by subsets of smoking status

| | n | minimum | maximum | median | mean | sd | p-value |
|---|---|---|---|---|---|---|---|
| Never | 135890 | -1.721 | 2.746 | 1.241 | 1.198 | 0.559 | - |
| Ever | 110348 | -1.733 | 2.835 | 1.241 | 1.196 | 0.561 | 0.4511 |
| “Occasionally” or “Once or twice” | 96272 | -1.731 | 2.927 | 1.249 | 1.207 | 0.555 | 0.0009 |
| NA | 1152 | -0.707 | 2.489 | 1.273 | 1.223 | 0.557 | 0.0945 |

*n*, number of observations; *sd*, standard deviation; *NA*, missing. The p-values are from Mann-Whitney U tests comparing the GS distributions to the Never group.

**Table S2.** FINEMAP top configuration of causal SNPs for the NMR on the chromosome 19 locus in the Finnish data

| SNP | BP | EA/NEA | MINOR | MAF | SNP PROB | BETAJ | SEJ | P-VALUEJ |
|---|---|---|---|---|---|---|---|---|
| RS12985907 | 41343544 | A/G | A | 0.28 | 0.9998 | -0.8 | 0.03 | $1.21 \times 10^{-118}$ |
| RS189621498 | 41288136 | A/G | A | 0.03 | 0.9763 | 0.61 | 0.08 | $9.9 \times 10^{-16}$ |
| RS1801272 | 41354533 | T/A | T | 0.02 | 0.9749 | -1.01 | 0.08 | $6.13 \times 10^{-34}$ |
| RS34945948 | 41340842 | G/A | G | 0.14 | 0.519 | 1.03 | 0.07 | $1 \times 10^{-54}$ |
| RS7248187 | 41437426 | C/G | G | 0.24 | 0.4319 | 0.32 | 0.03 | $4.63 \times 10^{-22}$ |
| RS116382863 | 41534881 | T/C | T | 0.13 | 0.1694 | 0.22 | 0.04 | $3.2 \times 10^{-9}$ |
| RS11466310 | 41861858 | T/C | T | 0.02 | 0.0714 | -0.54 | 0.1 | $4.18 \times 10^{-8}$ |
| RS7250713 | 41355195 | C/G | G | 0.37 | 0.028 | 0.33 | 0.03 | $1.99 \times 10^{-21}$ |
| RS74719953 | 41335799 | T/C | T | 0.09 | 0.0027 | -0.36 | 0.07 | $8.23 \times 10^{-8}$ |

FINEMAP results of the 5Mb region centered at the top SNP when only including SNPs passing quality control in UKB (n(SNP) = 10,133). The most probable configuration consisted of 9 SNPs (depicted in the table) and their heritability estimate was 32.1% (95% CI: 28.5–35.6%). FINEMAP gave a regional heritability estimate of 33.8% (95% CI: 30.0–37.8%) and suggested that there are 7–11 causal SNPs within the region.
*SNP*, single-nucleotide polymorphism; *BP*, base pair position in GRCh37 coordinates; *EA/NEA*, the effect allele/ the non-effect allele; *MINOR*, the less common allele; *MAF*, minor allele frequency in the Finnish dataset (n = 2,119) used for the FINEMAP analysis; *SNP PROB*, posterior probability of being a causal SNP; *BETAJ*, effect estimate from the joint model including all these 9 SNPs (reported for the effect allele); *SEJ*, standard error for the BETAJ; *P-VALUEJ*, p-value for the BETAJ.

**Table S4.** Linear regression beta coefficient for the zGS when explaining CPD by zGS in UKB

| Data | Number of observations | Beta coefficient | p-value |
|---|---|---|---|
| ALL | 23,682 | 0.07516 | $< 2 \times 10^{-16}$ |
| LOW GS ($zGS < 0$) | 10,871 | 0.10982 | $< 2 \times 10^{-16}$ |
| HIGH GS ($zGS \geq 0$) | 12,811 | 0.04237 | 0.0235 |

Analysis was done for the current smokers subset of UKB. CPD has been adjusted for sex, age and the first 10 genetic principal components, and then inverse normalized (rank-based-inverse-normal-transformation) before regressing it on zGS. The standard deviation of the adjusted CPD was 8.3. When looking at all individuals, each standard deviation increase in the GS is associated with a 0.07516 standard deviation increase in CPD, i.e. a 0.6 increase in cigarettes smoked per day. The regression was ran for three subsets of the data including either all individuals (ALL), only those with lower zGS values (LOW GS; $zGS < 0$) or only those with higher zGS values (HIGH GS; $zGS \geq 0$).

**Table S8.** Reason for stopping smoking by nicotine metabolism group (grouped based on the tertiles of the GS for the NMR)

| | Slow (n = 36,783) | Medium (n = 36,782) | Fast (n = 36,783) | p-value (slow vs fast) |
|---|---|---|---|---|
| Illness or ill health | 9.02 % | 8.76 % | 9.24 % | 0.3243 |
| Doctor's advice | 5.38 % | 5.35 % | 5.50 % | 0.5051 |
| Health precaution | 47.22 % | 47.86 % | 48.00 % | 0.0360* |
| Financial reasons | 18.36 % | 18.97 % | 19.24 % | 0.0024* |

Ever smokers who had stopped smoking for over 6 months during the time they smoked were asked: "Why did you stop smoking? (You can select more than one answer)" (UK Field 6157). Total number of ever smokers included in this analysis was $n = 110,348$, of which 26 % were current smokers and 74 % former smokers. *, Statistically significant difference ($p < 0.05$, 2-sample test for equality of proportions) between the Slow and Fast groups.

**Table S11.** Information on the NMR top SNP, rs56113850, and smoking status by ancestry group

| Data | rs56113850: C (vs T) frequency | HW exact p-value | info | missing proportion | n | Smoking Status (%) Never/Previous/Current/NA |
|---|---|---|---|---|---|---|
| White British | 0.57763 | 0.009 | 0.995 | 5.528e-10 | 343695 | 54.4 / 35.1 / 10.1 / 0.3 |
| Mixed | 0.511039 | 0.241 | 0.980 | 1.430e-08 | 2797 | 48.2 / 32.5 / 18.9 / 0.4 |
| Asian | 0.423935 | 8.525e-10 | 0.973 | 7.467e-09 | 9375 | 76.4 / 13.1 / 9.4 / 1.1 |
| Black | 0.388393 | 4.579e-05 | 0.972 | 1.313e-09 | 7618 | 69.7 / 17.2 / 12.3 / 0.8 |
| Chinese | 0.397583 | 0.001 | 0.900 | 5.988e-08 | 1503 | 78.8 / 13.3 / 7.6 / 0.3 |
| Other ethnic group | 0.483887 | 2.682e-10 | 0.965 | 6.890e-09 | 4354 | 60.7 / 25.3 / 13.3 / 0.7 |

$n$, number of observations; $NA$, missing.

### 3 Extended Content

#### 3.1 FINEMAP analyses

As in our previous study [1], we used the datasets YFS and FINRISK (n = 2,119), and the software tool FINEMAP (version 1.4), for rerunning the fine-mapping analyses of the chromosome 19 locus. We used a subset of the GWAS summary statistics and SNP correlation data from our previous FINEMAP analyses, which included SNPs within the ±2.5Mb flanking region of the top associating SNP (rs56113850). We included SNPs passing the following quality control criteria in both the Finnish and the UKB data: imputation info score > 0.7, call rate > 0.9, and Hardy-Weinberg Equilibrium $p > 10^{-6}$. Additionally, only SNPs with minor allele frequency (MAF) > 1 % in the Finnish data were included and multiallelic SNPs were excluded. Altogether, 10,133 SNPs were included in the FINEMAP analysis as opposed to the 12,060 SNPs included previously. We set the maximum number of allowed causal SNPs to 20, and otherwise used the default settings in FINEMAP.

#### 3.2 The GxE MR-pheWAS approach

The method has been described in detail by Millard et al. [2]. In short, mendelian randomization (MR) uses a genetic instrument as a proxy for an exposure in order to test for a causal effect of the exposure on the outcome, while avoiding bias due to confounding or reverse causality. The key assumption is that the genetic instrument has an association with the outcome solely through the exposure. Most approaches available for testing whether there is evidence for horizontal pleiotropy, and thus, violation of this assumption, are only applicable when several independent genetic instruments proxying the exposure have been used. However, when multiple genetic instruments are not available, such as in our case where our genetic instrument is reliant on only two association loci, the gene-by-environment (GxE) design, can be used.

The idea in GxE MR is to divide the data in to groups with different levels of the exposure, in our case, ever and never smokers. We assume that the effect of nicotine metabolism only occurs in people who are actually using nicotine, in other words our ever smokers subset. Then, if a given association between our genetic instrument and outcome is only due to nicotine metabolism, either directly or indirectly through traits such as amount smoked, we should only see an association among ever smokers, but not among never smokers. The approach permits us to distinguish whether the associations reflect a causal pathway through a) the NMR, either directly or through other traits such as amount smoked (effect only seen in ever smokers), b) some other pathways not including the NMR (same effect seen also in never smokers), or c) both (effect only seen in never smokers, or there are quantitative or directional differences in the effect sizes between ever and never smokers) (see Figure 1).
